# Bad company? The pericardium microbiome in people investigated for tuberculosis pericarditis in an HIV-prevalent setting

**DOI:** 10.1101/2024.04.26.24306431

**Authors:** Georgina Nyawo, Charissa Naidoo, Benjamin G. Wu, Benjamin Kwok, Jose C. Clemente, Yonghua Li, Stephanie Minnies, Byron Reeve, Suventha Moodley, Thadathilankal-Jess John, Sumanth Karamchand, Shivani Singh, Alfonso Pecararo, Anton Doubell, Charles Kyriakakis, Robin Warren, Leopoldo N. Segal, Grant Theron

## Abstract

**Background:** The microbiome likely plays a role in tuberculosis (TB) pathogenesis. We evaluated the site-of-disease microbiome and predicted metagenome in people with presumptive tuberculous pericarditis, a major cause of mortality, and explored for the first time, the interaction between its association with C-reactive protein (CRP), a potential diagnostic biomarker and the site-of-disease microbiome in extrapulmonary TB.

**Methods:** People with effusions requiring diagnostic pericardiocentesis (n=139) provided background sampling controls and pericardial fluid (PF) for 16S rRNA gene sequencing analysed using QIIME2 and PICRUSt2. Blood was collected to measure CRP.

**Results:** PF from people with definite (dTB, n=91), probable (pTB, n=25), and non- (nTB, n=23) tuberculous pericarditis differed in β-diversity. dTBs were, vs. nTBs, *Mycobacterium-, Lacticigenium-,* and *Kocuria-*enriched. Within dTBs, HIV-positives were *Mycobacterium-, Bifidobacterium-*, *Methylobacterium-*, and *Leptothrix*-enriched vs. HIV-negatives and HIV-positive dTBs on ART were *Mycobacterium*- and *Bifidobacterium*-depleted vs. those not on ART. Compared to nTBs, dTBs exhibited short-chain fatty acid (SCFA) and mycobacterial metabolism microbial pathway enrichment. People with additional non-pericardial involvement had differentially PF taxa (e.g., *Mycobacterium*-enrichment and *Streptococcus*-depletion associated with pulmonary infiltrates). *Mycobacterium* reads were in 34% (31/91), 8% (2/25) and 17% (4/23) of dTBs, pTBs, and nTBs, respectively. β-diversity differed between patients with CRP above vs. below the median value (*Pseudomonas*-depleted). There was no correlation between enriched taxa in dTBs and CRP.

**Conclusions:** PF is compositionally distinct based on TB status, HIV (and ART) status and dTBs are enriched in SCFA-associated taxa. The clinical significance of these findings, including mycobacterial reads in nTBs and pTBs, requires evaluation.

## Introduction

The burden of tuberculosis (TB) is severe in sub-Saharan Africa, where HIV is frequent^1^. In 2021, South Africa reported >300 000 incident TB cases and 61 000 deaths^1^. About 16% of incident cases are extrapulmonary, affecting sites outside the lungs^2^. Extrapulmonary TB (EPTB) has high fatality, especially in people living with HIV (PLHIV)^3^. Tuberculous pericarditis (TBP) is an especially severe manifestation, and the deadliest manifestation of EPTB, with up to 40% mortality^4-6^.

Immune responses to *Mycobacterium tuberculosis* complex (*Mtb*) involve a cascade of events that are, in part, influenced by the microbiome and their metabolites^7^ ^8^. For instance, gut enrichment with specific anaerobes (*Anaerostipes, Blautia, Erysipelotrichaceae*) independently predicts activation of host inflammatory immune pathways and active pulmonary TB compared to sick controls^9^ ^10^. In PLHIV on ART, greater microbially-derived short chain fatty acid (SCFA; butyrate, propionate) concentrations in the lower airways independently predict incident TB^11^. When SCFAs are added at physiological concentrations to alveolar macrophages *ex vivo*, there are reductions in macrophage-mediated *Mtb* kill^11^.

C-reactive protein (CRP) is a non-specific blood inflammatory marker for triage^12^, especially in PLHIV in primary care, where it is WHO-recommended^13^. In studies where CRP was co-analysed with the microbiome, CRP negatively correlated with abundances of anti-inflammatory-associated gut microbiota^14^ such as *Faecalibacterium^15^, Lactobacillus* and *Bifidobacterium^16^*. In COVID-19 patients, CRP correlated with potentially pathogenic bacteria such as *Klebsiella* spp. and *Enterococcus* spp. ^17^ However, the relationship between CRP and the site-of-disease microbiota in presumptive TBP has not been studied.

Little is known about the microbiome of the pericardial space, especially in the context of EPTB, a major cause of pericardial disease. Previous research has hinted at the influence of microbiota on TB, making it a promising treatment avenue^7-9^. Our work showed the presence of distinctive microbial communities at the site of disease in people with presumptive tuberculous lymphadenitis^18^. Importantly, these communities differed by TB status (definite-, probable- or non-TB) and clinical characteristics, with specific community states (i.e., *Mycobacterium*-dominated lymphotype) being associated with more severe disease^18^.

Together, this can aid in the identification of novel biomarkers at the microbial-host interface that leads to strategies like host-directed therapies for TB prevention and treatment, including in PLHIV. This could also enhance our understanding of severe inflammation in EPTB, which partly contributes to its high morbidity and mortality. Understanding interactions in the microbiome, together with correlations with CRP can help find meaningful networks via which microbes and host interact, and this can help uncover novel associations and mechanisms.

We therefore evaluated the site-of-disease microbiome and its predicted metagenome in people investigated for TBP (including PLHIV) as well as whether CRP as a surrogate for inflammation correlates with specific taxa. This work will help provide foundational data on the microbiome’s role in TBP.

## Methods

### Recruitment and follow-up

People with presumed TB pericarditis referred to Tygerberg Academic Hospital in Cape Town, South Africa for routine diagnostic investigation including pericardiocentesis were recruited between 24 November 2016 and 9 April 2021. Eligible people were ≥18 years old and not on TB treatment for ≥2 weeks. Demographic, clinical, and chest X-ray (CXR) data were collected. People with TBP were managed by the overseeing clinician in line with current guidelines for the management and treatment of pericardial effusions, and assessed by telephonic follow-up after a minimum of 12 weeks. The study had no role in patient management.

### Ethics

All people provided written informed consent. The study was approved by the Stellenbosch University Health Research and Ethical Committee (HREC, N16/04/050), Tygerberg Academic Hospital, and the Western Cape Department of Health (WC_2016RP15_762).

### Specimen collection and processing

Two sampling controls were collected per person: a skin swab of the sterilised puncture site and a sterile saline (Ysterplaat Medical Supplies, South Africa) flush of the vascular sheath prior to pericardiocentesis (background control). At least 4mL pericardial fluid (PF) was collected once sufficient volumes were obtained for routine diagnostic testing. Samples were stored at −80°C until batched DNA extraction. Peripheral blood was collected in serum-separating tubes (SST tubes; BD), and centrifuged (2054*g*, 10 min) within four hours of collection, and serum stored at −80°C. Further details on specimen collection and microbiological testing is in the **Supplementary material**.

### Patient classification

We designated people as definite tuberculous pericarditis (dTB), probable tuberculous pericarditis (pTB), or non-tuberculous pericarditis (nTB) per **Table S1**^19^. People were classified as dTBs if they were positive for acid-fast bacilli by Ziehl-Neelsen smear microscopy and/or had at least one MTB-positive specimen by Mycobacteria Growth Indicator Tube (MGIT) 960 liquid culture (BD Diagnostics, USA), Xpert MTB/RIF (Xpert; Cepheid, USA) and/or Xpert MTB/RIF Ultra (Ultra; Cepheid, USA). pTBs did not meet dTB criteria, had no alternative diagnosis, and were empirically prescribed TB therapy. nTBs had no microbiological evidence of TB and were not placed on treatment.

### Laboratory procedures

The PureLink Microbiome DNA Purification Kit (Invitrogen, USA) was used for PF, skin swabs and, for efficiency, 1 in 5 background samples. The 16S rRNA gene V4 hypervariable region (150 bp read length, paired-end) was amplified and underwent Illumina MiSeq sequencing^11^. Further detail is in the **Supplementary Material**. CRP was measured in bio-banked serum using the Cobas High-Sensitivity Immunoturbidimetric Assay (Roche Diagnostics Limited, Burgess Hill, UK) per the manufacturer’s <u>instructions</u>.

### Microbiota analyses

#### Pre-processing

Sequences were pre-processed using QIIME (version 2020.8)^20^ and DADA2^21^. Amplicon sequence variants (ASVs) were assigned at 99% similarity against representative sequences in Greengenes (version 13.8)^22^ with zero abundance taxa removed.

#### Taxonomic analyses

α- and β-diversities were calculated using *vegan* (v2.6-4)^23^. Differential taxa and pathway abundance analyses used *DESeq2* (v1.22.2)^24^ with Benjamini– Hochberg (BH) multiple testing correction after feature tables were pruned (genera not seen >5 times in ≥1% of samples removed)^25^. CRP was categorised as high or low if the concentration readout was above or below the median for the people included in that analysis. Detailed methods for identifying potential contaminants and clustering analysis are in the **Supplementary Material.**

#### Functional pathway analyses

Phylogenetic Investigation of Communities by Reconstruction of Unobserved States (PICRUSt2; v.2.1.3-b)^26^ was used for metagenomic inference with default parameters (picrust2_pipeline.py) and metabolic function predictions based on MetaCyc^27^.

#### Correlation analysis with CRP and microbiome

Only sequence data with paired CRP measurements were included. Microbial relative abundances were correlated with CRP measurements using Correlations Under the Influence (CUTIE)^28^.

### Statistical analyses

Analyses used GraphPad Prism (v7; GraphPad Software, USA), STATA (v16; StataCorp, USA) and R (v4.2; R Core Team). Medians with interquartile ranges (IQRs) were reported for continuous variables, and the number and percentage were reported for categorical variables. Non-parametric tests (microbiome data are not normally distributed^29^) included the Mann-Whitney or Wilcoxon signed-rank test for unpaired and paired comparisons involving two groups, Kruskal-Wallis or Friedman tests for unpaired and paired comparisons involving ≥2 groups, and Spearman’s Rho for correlation analyses. Permutational multivariate analysis of variance (PERMANOVA) was computed with 999 permutations for β-diversity, and *R*^2^ used to measure variation explained by a variable. The Benjamini–Hochberg procedure was used to correct for multiple comparisons by controlling the false discovery rate (FDR)^25^. Adjusted *p*-values of <0.2 and <0.05 were considered significant for taxa and pathways, respectively^30^. More information is in the **Supplementary Methods**.

## Results

### Cohort characteristics

Our cohort was comprised of 91 (65%), 25 (18%) and 23 (17%) dTBs, pTBs, and nTBs, respectively (**Table 1)**. Compared to nTBs, dTBs were more likely to have HIV, cough, weight loss, be on medication (other than TB medication) and, in their PF, have lower albumin but higher adenosine deaminase (ADA), lactate dehydrogenase (LDH), and unstimulated interferon-gamma (uIFN-γ) concentrations. pTBs were more likely than nTBs to have HIV, previous TB, and TB symptoms and, in PF, lower albumin and higher ADA and uIFN-γ concentrations.

**Table 1:** Demographic and clinical characteristics of people with presumptive TB pericarditis. dTBs were more likely to have HIV, cough, weigh less, be using non-TB medications and, in PF, have lower albumin concentrations and higher ADA, LDH, and uIFN-γ concentrations than nTBs. pTBs were more likely to have HIV, previous TB, more self-reported symptoms, have lower albumin concentration, and have higher ADA and uIFN-γ fluid concentration, than nTBs. dTBs were less likely to already be on treatment at recruitment (< 2 weeks) compared to pTBs. Data are n/N (%) or median (IQR).

|  | People with presumptive TB |  |  |  | *p-value* |  |  |
| --- | --- | --- | --- | --- | --- | --- | --- |
|  | Total (n=139) | dTB (n=91) | pTB (n=25) | nTB (n=23) | dTB vs. pTB | dTB vs. nTB | pTB vs. nTB |
| Age, years | 37 (31, 48) | 40 (31, 48) | 34 (31, 46) | 37 (31, 58) | 0.910 | 0.563 | 0.485 |
| Sex (Female) | 59/139 (42) | 37/91 (41) | 10/25 (40) | 12/23 (52) | >0.999 | 0.450 | 0.580 |
| Current tobacco smoker | 30/138 (22) | 17/90 (19) | 6/25 (24) | 7/23 (30) | 0.780 | 0.360 | 0.860 |
| Previous tobacco smoker | 29/108 (27) | 19/73 (26) | 7/19 (37) | 3/16 (19) | 0.370 | 0.950 | 0.420 |
| Clinical characteristics: |  |  |  |  |  |  |  |
| HIV | 75/139 (54) | 53/91 (58) | 20/25 (80) | 2/23 (9) | **0.046** | **<0.0001** | **<0.0001** |
| On ART | 26/73 (37) | 18/53 (34) | 7/18 (39) | 1/2 (50) | 0.930 | >0.999 | >0.999 |
| CD4 count, cells/μl | 158 (54, 319) | 151 (49, 328) | 208 (97, 339) | 106 (36, 176) | >0.999 | >0.999 | 0.971 |
| Previous TB | 31/136 (23) | 21/88 (24) | 9/25 (36) | 1/23 (4) | 0.339 | 0.072 | **0.019** |
| Self-reported symptoms: |  |  |  |  |  |  |  |
| Cough | 76/132 (58) | 52/85 (61) | 16/24 (67) | 8/23 (35) | 0.801 | **0.043** | 0.058 |
| Fever | 52/131 (40) | 35/85 (41) | 12/23 (52) | 5/23 (22) | 0.480 | 0.142 | 0.067 |
| Night sweats | 68/133 (51) | 45/86 (52) | 15/24 (63) | 8/23 (35) | 0.510 | 0.210 | 0.110 |
| Weight loss | 85/134 (63) | 60/87 (69) | 17/24 (71) | 8/23 (35) | >0.999 | **0.006** | **0.029** |
| Chest X-ray results: |  |  |  |  |  |  |  |
| Cardiomegaly | 93/139 (67) | 61/91 (67) | 17/25 (68) | 15/23 (65) | >0.999 | >0.999 | >0.999 |
| Pulmonary infiltrates | 31/139 (22) | 23/91 (25) | 4/25 (16) | 4/23 (17) | 0.480 | 0.600 | >0.999 |
| Hilar lymphadenopathy | 11/139 (8) | 7/91 (8) | 2/25 (8) | 2/23 (9) | >0.999 | >0.999 | >0.999 |
| Miliary pattern | 7/139 (5) | 6/91 (7) | 1/25 (4) | 0/23 (0) | 0.990 | 0.460 | >0.999 |
| Pleural effusion | 88/139 (63) | 60/91 (66) | 16/25 (64) | 12/23 (52) | >0.999 | 0.330 | 0.590 |
| Pericardial tamponade | 47/127 (37) | 29/81 (36) | 9/23 (39) | 9/23 (39) | 0.960 | 0.960 | >0.999 |
| Treatment history: |  |  |  |  |  |  |  |
| Current TB treatment (<2 weeks) | 21/139 (15) | 12/91 (13) | 9/25 (36) | 0/23 (0) | **0.020** | 0.144 | **0.005** |
| No TB treatment | 118/139 (85) | 79/91 (87) | 16/25 (64) | 23/23 (100) | **0.020** | 0.144 | **0.005** |
| Other medications | 58/129 (45) | 33/83 (40) | 9/23 (39) | 16/23 (70) | >0.999 | **0.210** | 0.076 |
| Non-TB antibiotic use within 1 year | 21/125 (17) | 12/80 (15) | 7/24 (29) | 2/17 (12) | 0.200 | >0.999 | 0.350 |
| Current non-TB antibiotic use | 17/21 (81) | 10/12 (83) | 5/7 (71) | 2/2 (100) | 0.980 | >0.999 | >0.999 |
| Total volume aspirated (mL) | 700 (500, 1000) | 670 (500, 915) | 720 (500, 1000) | 750 (400, 1050) | >0.999 | >0.999 | >0.999 |
| Fluid cytochemistry: |  |  |  |  |  |  |  |
| ADA (U/L) | 48 (26, 67) | 51 (34, 68) | 50 (38, 65) | 16 (11, 33) | >0.999 | **0.001** | **0.016** |
| Protein (g/L) | 58 (50, 65) | 58 (49, 64) | 61 (55, 67) | 60 (50, 64) | >0.999 | >0.999 | >0.999 |
| LDH (g/L) | 696 (424, 1440) | 840 (451, 1440) | 645 (541, 1490) | 408 (181, 842) | >0.999 | **0.016** | 0.132 |
| Albumin (U/L) | 22 (16, 26) | 21 (16, 25) | 23 (19, 26) | 28 (25, 30) | >0.999 | **0.010** | 0.126 |
| uIFN-γ (pg/mL) | 803 (3, 2510) | 1150 (165, 2590) | 885 (4, 2810) | 28 (0, 5) | >0.999 | **<0.0001** | **<0.0001** |
| Blood CRP (mg/L)* | 125 (71, 177) | 137 (75, 176) | 136 (93, 202) | 96 (45, 174) | >0.999 | >0.999 | >0.999 |
| Mortality | 24 (18) | 14/88 (16) | 4/24 (17) | 6/22 (27) | >0.999 | 0.350 | 0.610 |

\*For CRP, paired samples were available from 112/139 participants (72/91 dTB; 20/25 pTB and 20/23 nTB). Missing data:
Current tobacco smoker (n=1), Previous tobacco smoker (n=31), ART status (n=2), Previous TB (n=3), Self-reported symptoms
(n=8), Other medications (n=10), Non-TB antibiotic use within 1 year (n=14). Bold indicates statistical significance (P < 0.05).
Abbreviations: ADA: adenosine deaminase; ART: antiretroviral therapy; CRP: C-reactive protein; dTB: definite tuberculous
pericarditis; IQR: interquartile range; LDH: lactate dehydrogenase; nTB: non-tuberculous pericarditis; pTB: probable
tuberculous pericarditis; TB: tuberculosis; TBP: tuberculous pericarditis; and uIFN-γ : unstimulated interferon gamma

### Pericardial fluid shares few taxa with skin and background controls

PF α-diversity was lower than background and skin. PF β-diversity differences occurred with *Mycobacterium-* (vs. skin and background) and *Streptococcus-*enrichment (vs. skin only) (**Figure S1A-D**). None of the top four potentially contaminating taxa identified appeared in differential analyses (**Figures S2-3**, **<u>additional table</u>**).

### Mycobacterium and other potential opportunistic pathogens are enriched in dTBs

#### Differences by TB status

We did not detect α-diversity differences (**Figure 2A**). β-diversity differed between dTBs and pTBs, as well as pTBs vs. nTBs (PERMANOVA *p*=0.020 and 0.003, respectively, **Figure 2B**), but not between dTBs and nTBs. dTBs were *Mycobacterium-*, *Lacticigenium-*, and *Kocuria-*enriched vs. nTBs (**Figure 2C**), and *Mycobacterium-*, *Lacticigenium-* and *Caulobacter-*enriched and *Streptococcus*-depleted vs. pTBs (**Figure 2D**). pTBs were *Streptococcus-* (vs. dTBs) and *Novosphingobium-*enriched vs. nTBs (**Figure 2D-E**), and more phylogenetically dissimilar to each other. dTBs were the most phylogenetically similar to each other followed by nTBs (**Figure S4**).

**Figure 1:**
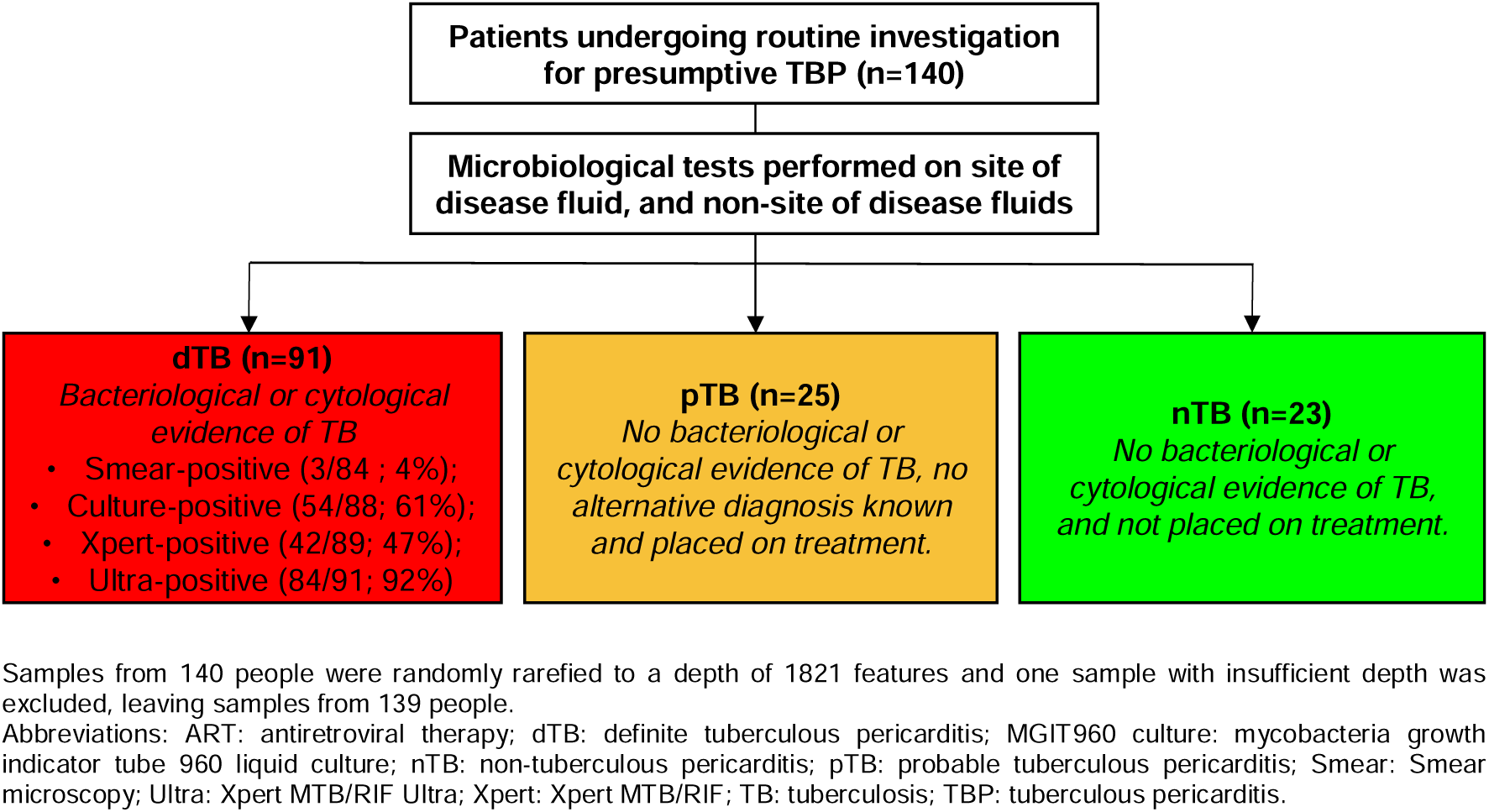
Study flow chart. Pericardial fluid, skin and background controls were collected from 140 presumptive TBP people. dTBs: definite tuberculous pericarditis; nTBs: non-tuberculous pericarditis; pTBs: probable tuberculous pericarditis; Smear: Smear microscopy; MGIT960 Culture: Mycobacteria Growth Indicator Tube 960 liquid culture; Xpert: Xpert MTB/RIF; Ultra: Xpert MTB/RIF Ultra.

**Figure 2:**
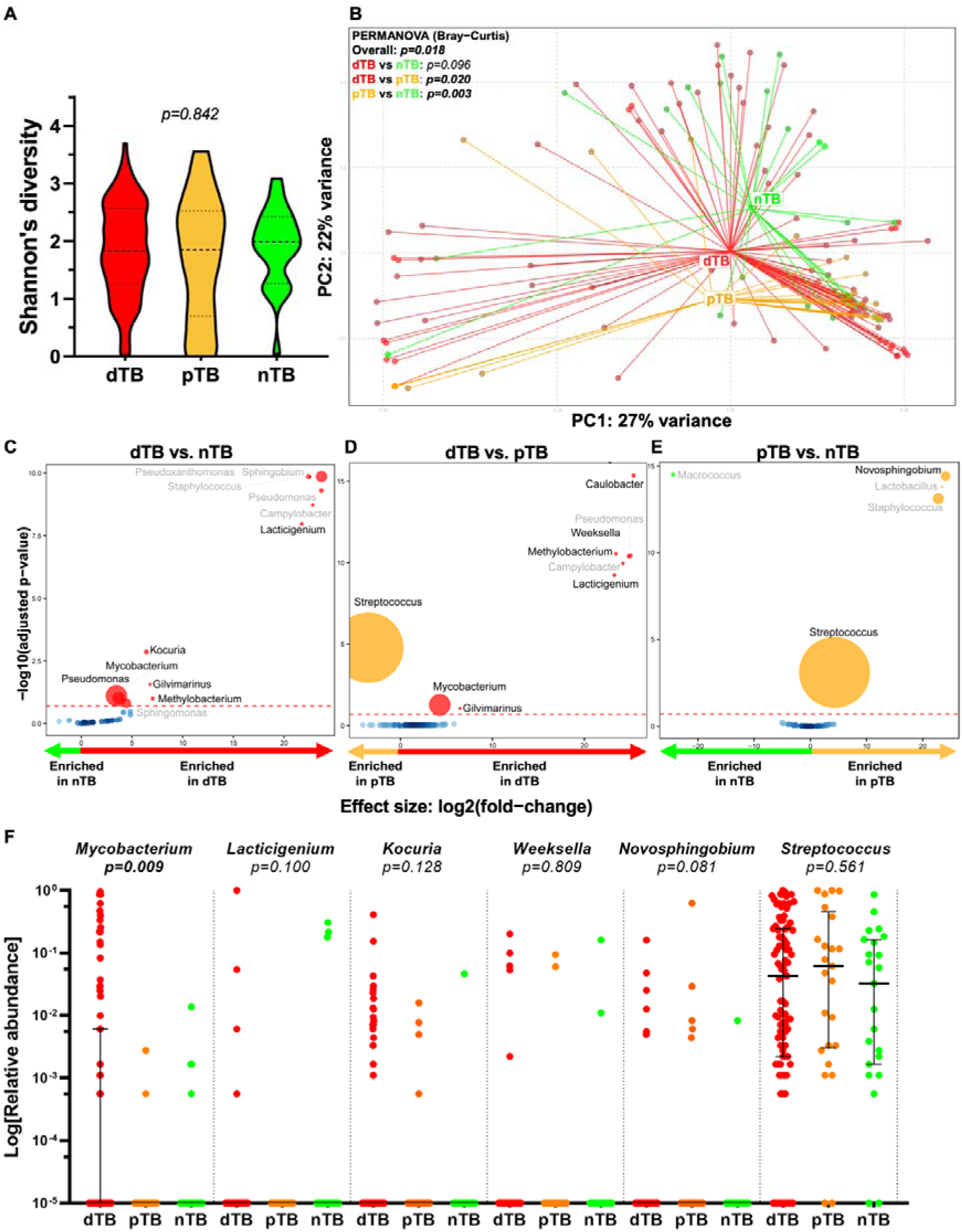
dTBs have a distinct microbiome enriched in *Mycobacterium* and other potential opportunistic pathogens. **(A)** Although α-diversity, which measures species diversity, was similar across the three groups, **(B)** β-diversity, which measures differences in microbial composition, differed significantly between PF microbiota of pTBs vs. dTBs and pTBs vs. nTBs. **(C)** dTBs vs. nTBs were *Mycobacterium-*, *Lacticigenium-* and *Kocuria*-enriched, and *Streptococcus*-depleted. **(D)** dTBs vs. pTBs were *Mycobacterium-*, *Lacticigenium-*, and *Caulobacter*-enriched and *Streptococcus*-depleted. **(E)** pTBs vs. nTBs were *Novosphingobium-* and *Streptococcus-enriched* (red dotted line represents adjusted *p*-value=0.2; circle size represents relative abundance; taxa identified as possible contaminants are shown in grey). **(F)** Genus-level average relative abundances of top taxa differentiating between TB groups. dTB: definite tuberculous pericarditis; nTB: non-tuberculous pericarditis; PF: pericardial fluid; pTB: probable tuberculous pericarditis; TB: tuberculosis; TBP: tuberculous pericarditis.

#### Grouping pTBs with dTBs or nTBs

When pTBs were grouped with dTBs or nTBs, there were no α- or β-diversity differences vs. nTBs or dTBs, respectively (**Figure S5A-D**). dTBs were *Mycobacterium-* and *Lacticigenium*-enriched vs. nTBs regardless of with whom pTBs were grouped (**Figure S5E-F**).

#### 16S rRNA gene sequencing correlates with diagnostic results

Mycobacterial reads occurred in more dTBs than pTBs [34% (31/91) vs. 8% (2/25), *p*=0.011] but did not reach significance when dTBs were compared to nTBs [17% (4/23), *p*=0.181] (**Figure 2F, Figure S6**). Mycobacterial reads inversely correlated with PCR (r=-0.361, *p*<0.012; **Figure S7A**) and culture measures of mycobacterial load (r=-0.484, *p*<0.001; **Figure S7B**).

### Bifidobacterium enrichment is associated with HIV

Although α- and β-diversity did not differ by HIV status (**Figure S8A-B**), PLHIV were *Mycobacterium*-enriched vs. HIV-negatives (**Figure S8C**).

#### Comparisons by HIV status within different TB groups

No α- and β-diversity differences by HIV status occurred in dTBs or pTBs (**Table S2**). In dTBs, HIV-positives were *Methylobacterium*-, *Bifidobacterium*-, and *Leptothrix*-enriched vs. HIV-negatives (**Figure S8D**). nTBs were not compared by HIV status due to a small sample size (n=2 HIV-positives).

#### Comparisons by TB status within people of the same HIV status

In HIV-positives, α- diversity was similar by TB status (**Figure 3A**) but β-diversity differed (*p*=0.038; **Figure 3B**) and dTBs were *Mycobacterium-, Bifidobacterium-,* and *Leptothrix-*enriched vs. pTBs (**Figure 3C**). α- and β-diversities were similar between HIV-positives with high vs. low CD4 counts (dichotomised at median value, data not shown), where those with lower counts were *Mycobacterium*-enriched vs. people with higher counts (**Figure S9A-B**). In HIV-negatives, α- and β-diversities were similar between TB groups (**Figure 3D-E**) and taxonomic differences were only observed between dTBs vs. pTBs, with dTBs *Kocuria-, Halomonas-,* and *Gilvimarinus*-enriched (**Figure 3F**).

**Figure 3:**
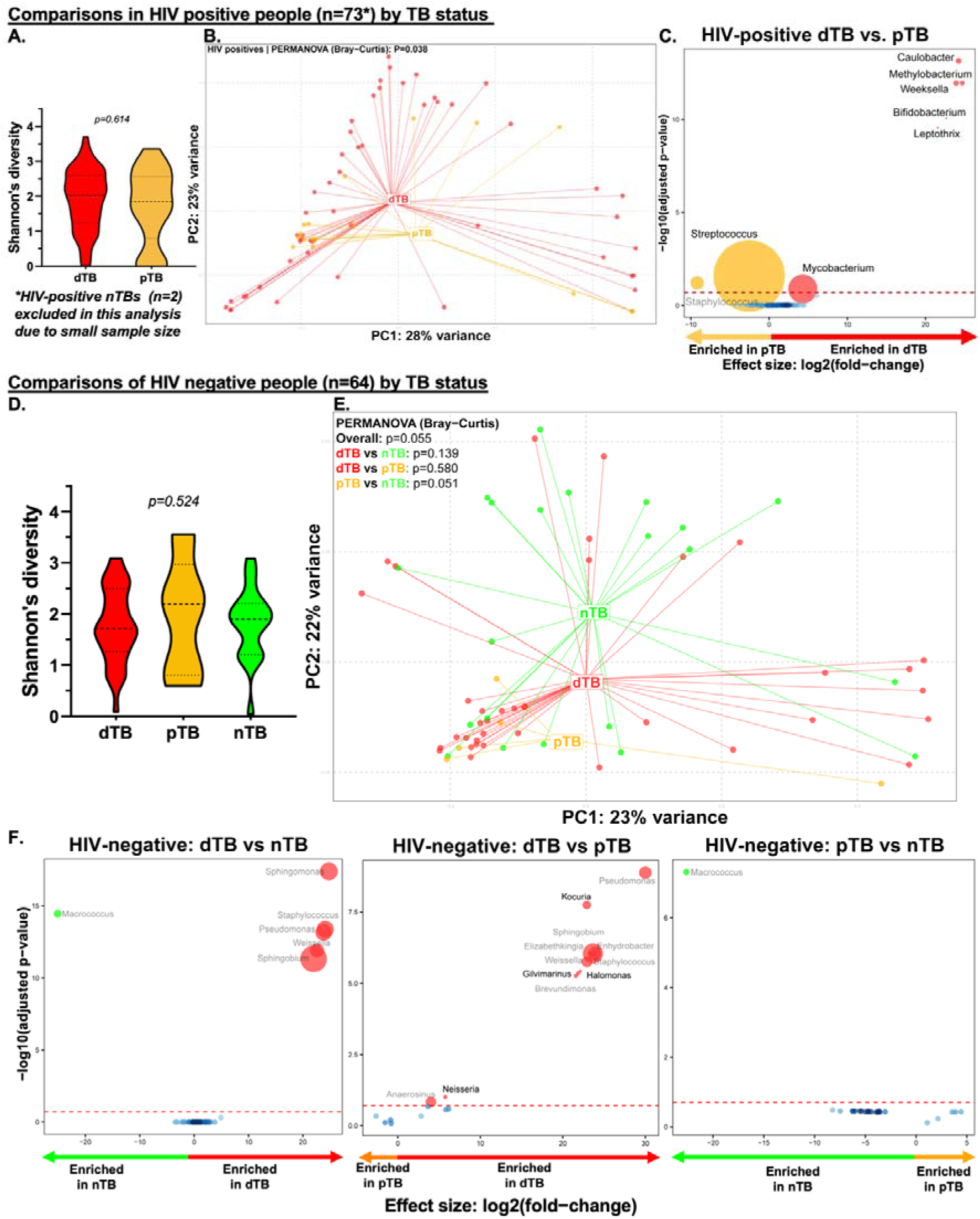
*Bifidobacterium* enrichment is associated with *Mycobacterium* and HIV positivity in TBP. In PLHIV, **(A)** α-diversity did not differ by TB status, **(B)** however, β- diversity did, **(C)** where HIV-positive dTBs were *Mycobacterium-, Bifidobacterium-,* and *Leptothrix-*enriched, and *Streptococcus-*depleted compared to HIV-positive pTBs. In people without HIV, both **(D)** α-diversity and **(F)** β-diversity did not differ by TB status, and HIV-negative dTBs were *Mycobacterium, Caulobacter,* and *Weeksella-*enriched, and *Streptococcus-*depleted compared to HIV-negative pTBs (red dotted line represents adjusted *p*-value=0.2; circle size represents relative abundance; taxa identified as possible contaminants are shown in grey). dTB: definite tuberculous pericarditis; HIV+: HIV-positive; HIV-: HIV-negative; nTB: non-tuberculous pericarditis; PF: pericardial fluid; pTB: probable tuberculous pericarditis; TB: tuberculosis; TBP: tuberculous pericarditis

#### Comparisons by ART status in PLHIV

A third of PLHIV were on ART (**Table 1**). α-Diversity was similar by ART status, but β-diversity differed (**Table S2**). Those on ART were *Streptococcus*-enriched and *Mycobacterium-* and *Bifidobacterium-*depleted vs. those not on ART (**Figure S9C**) and conclusions were similar when restricted to dTBs (**Figure S9D**).

### Chest abnormalities associated with the microbiome

We evaluated microbiome differences by CXR results (**Table 1**) and found no α-diversity differences. However, β-diversity differed in people with vs. without pulmonary infiltrates (PIs; **Table S2**), with people with PIs *Mycobacterium*-enriched and *Streptococcus*-depleted (**Figure 4A**). β-diversity also differed in people with or without pleural effusions (PEs) (**Table S2**), where people with PEs were *Streptococcus*- and *Pseudomonas*-enriched and *Brachymonas*-depleted (**Figure 4B**).

**Figure 4:**
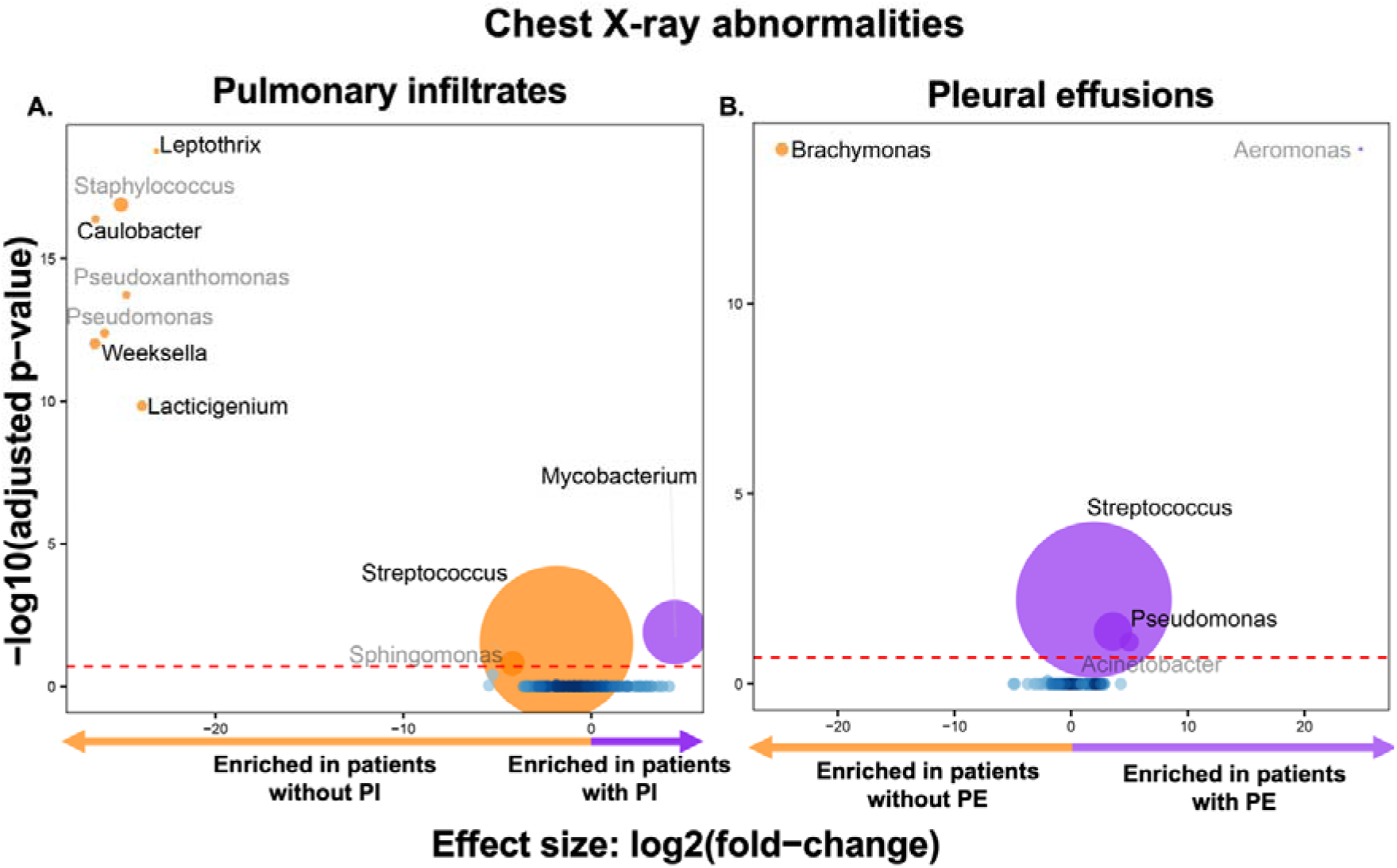
Correlations between imaging and clinical characteristics and the PF microbiota. Pulmonary infiltrates (PI) and pleural effusions (PE) are assotiated with PF microbiome. Compared to patients without PI, **(A)** presumptive TBPs with a PI were *Mycobacterium*-enriched and *Lacticigenium-* and *Streptococcus*-depleted, whilst **(B)** compared to patients without PE, presumptive TBP people with PE were *Streptococcus-* and *Pseudomonas-*enriched, and *Brachymonas*-depleted (red dotted line represents adjusted *p*-value=0.2; circle size represents relative abundance; taxa identified as possible contaminants are shown in grey). dTB: definite tuberculous pericarditis; PF: pericardial fluid; PE: pleural effusions; PI: pulmonary infiltrates; pTB: probable tuberculous pericarditis; TB: tuberculosis; TBP: tuberculous pericarditis

### People with definite TBP are enriched in SCFA metabolic pathways

Compared to nTBs, dTBs were enriched with SCFA metabolism (*L-lysine fermentation to acetate and butanoate*), aromatic compound degradation (*cinnamate and 3-hydroxycinnamate degradation to 2-oxopent-4-enoate*), and mycobacterial metabolism (*superpathway of mycolyl-arabinogalactan-peptidoglycan complex biosynthesis*) pathways (**Figure 5**). Amongst dTBs, HIV-positives were depleted in biosynthesis pathways [*GDP-D-glycero-*α*-D-manno-heptose biosynthesis*, *peptidoglycan biosynthesis V (*β*-lactam resistance)* and *superpathway of geranylgeranyl diphosphate biosynthesis II (via MEP)*] compared to HIV-negatives (**Figure S10**). Comparisons involving pTBs are in **Figures S11-S12.**

**Figure 5:**
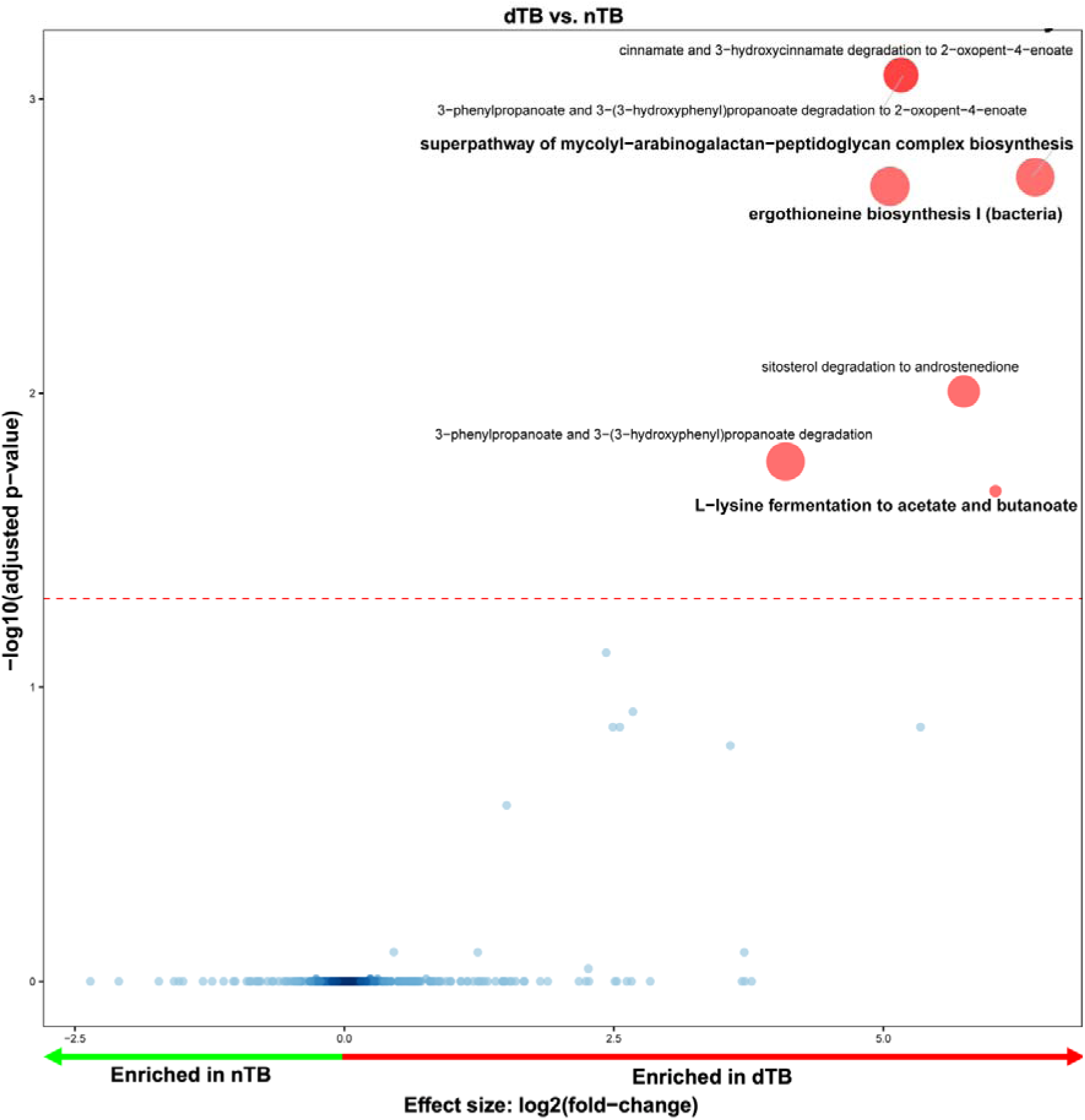
Inferred functional content shows enrichment of SCFA metabolic pathways in the PF of TBP. PF in dTBs infer a disctint functional capacity, including an enrichment of pathways associated with the production of short-chain fatty acids (SCFAs), aromatic compound degradation, and mycobacterial metabolism vs. nTBs (red dotted line represents adjusted *p*-value=0.05; circle size represents relative abundance; pathways of interest are bolded). dTB: definite tuberculous pericarditis; PF: pericardial fluid; nTB: non-tuberculous pericarditis; TB: tuberculosis; TBP: tuberculous pericarditis; SCFA: short chain fatty acids.

### The association between CRP and the microbiome

#### Microbial composition differed with *Pseudomonas* depleted in high vs. low CRP

CRP concentrations did not differ by TB status [median 137.30 (IQR 75.30 - 176.30) ng/mL in dTBs vs. 95.96 (IQR 45.25 - 173.70) ng/mL in nTBs] (**Figure 6A**) nor correlated with *Mycobacterium* reads (**Figure 6B-C**). People with above median CRP concentrations [median 125.4 (IQR70.83 – 176.7) ng/mL] had, vs. those below the median, no α-diversity differences (**Figure 6D**) but different β-diversity (**Figure 6E**), with *Pseudomonas-*depletion (**Figure 6F**). These differences were similar when restricted to people of a specific TB (**Table 2**) or HIV status (**Figures S13-14**), with a few additional taxa differentially enriched. The top three taxa enriched in dTBs (*Mycobacterium*, *Lacticigenium*, *Kocuria*) did not correlate with CRP levels (p-values of 0.523, 0.260, and 0.999, respectively).

**Figure 6:**
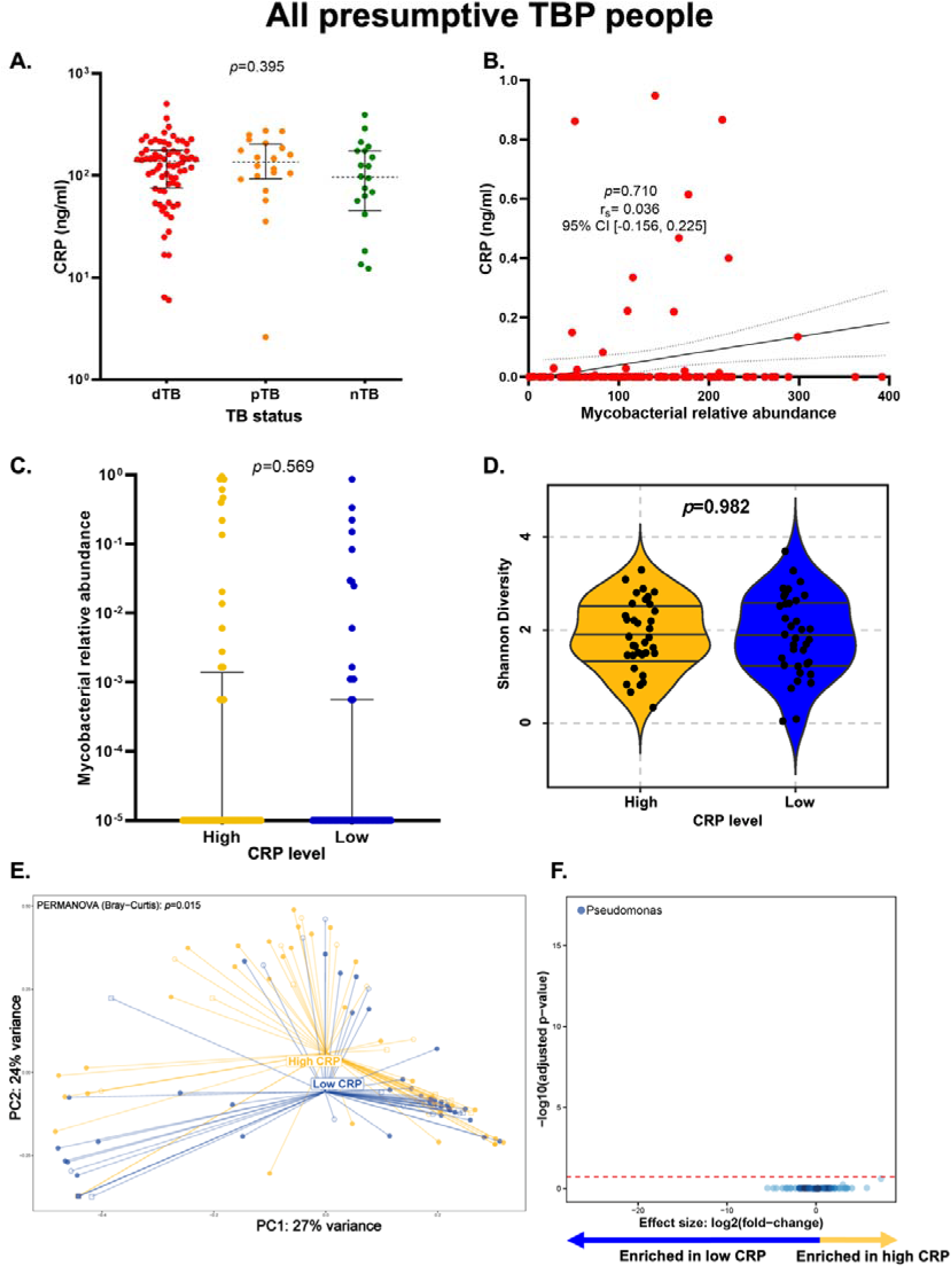
Microbial composition differs in presumptive TBP people with high vs low CRP. **A)** CRP levels are similar in dTBs vs. pTBs vs. nTBs. **B)** Mycobacterial reads do not correlate with CRP levels; r_s_ represents the Spearman’s Rho coefficient for the correlation of mycobacterial reads with CRP. People with high CRP levels have similar **C)** *Mycobacterium* reads and **D)** α-diversity in their pericardial fluid compared to people with low CRP levels (solid circles: dTB; empty squares: pTB; empty circles: nTB). **E)** β-diversity differs between people with high vs low CRP. **F)** *Pseudomonas* is depleted in people with high vs low CRP. Discriminatory taxa appear above the threshold (red dotted line, FDR=0.2); circle size represents relative abundance. CI, Confidence interval; CRP,LC-reactive protein; dTB: definite-TBP; FDR, false discovery rate; nTBP, non-TBP; PERMANOVA, permutational multivariate analysis of variance; pTB: probable-TBP; TBP: tuberculous pericarditis.

**Table 2:** CRP levels do not affect microbiome α-diversity, but influence β-diversity in people with presumptive TBP. α-diversity remained consistent regardless of CRP concentration, while significant differences in β-diversity were observed, indicating distinct microbial compositions between the two groups.

| High vs. low CRP | Overall (n=139) |  |  | dTb (n=91) |  |  | pTB (n=25) |  |  | nTB (n=23) |  |  |
| --- | --- | --- | --- | --- | --- | --- | --- | --- | --- | --- | --- | --- |
| | α-diversity | β-diversity | | α-diversity | β-diversity | | α-diversity | β-diversity | | α-diversity | β-diversity | |
|  |  | *p-value* | *R<sup>2</sup> value* |  | *p-value* | *R<sup>2</sup> value* |  | *p-value* | *R<sup>2</sup> value* |  | *p-value* | *R<sup>2</sup> value* |
| All participants | 0.982 | **0.015** | 0.02723 | 0.982 | **0.013** | 0.02723 | 0.2265 | 0.527 | 0.0461 | 0.5967 | 0.3333 | 0.04407 |
| Within HIV-positives | 0.8932 | 0.059 | 0.02537 | 0.5028 | 0.05 | 0.03891 | 0.6744 | 0.826 | 0.04857 | 0.3173 | - | - |
| Within HIV-negatives | 0.1432 | 0.526 | 0.01987 | 0.2282 | 0.719 | 0.03419 | 0.1213 | 0.3333 | 0.36901 | 0.6911 | 0.668 | 0.04903 |

Bold indicates statistical significance ( $P < 0.05$ ). Abbreviations: CRP: C-reactive protein; dTB: definite tuberculous pericarditis; nTB: non-tuberculous pericarditis; pTB: probable tuberculous pericarditis; TB: tuberculosis.

## Discussion

Our key findings are that 1) compared to PF from people without TBP, PF from people with TBP differs both in terms of β-diversity and the relative abundance of specific taxa, including those other than *Mycobacterium* (*Lacticigenium*, *Kocuria*, and *Weeksella*), 2) HIV is associated with the differential abundance of *Mycobacterium* overall, and within dTBs, *Bifidobacterium* is associated with proxies of more severe disease (HIV-positivity and, in PLHIV, no ART), and 3) taxonomic differences between dTBs and nTBs manifest in different functional capacities, including enrichment of SCFA production pathways. Furthermore, 4) the PF microbiome is associated with distinctive clinical and chest imaging characteristics and 5) many people with dTB do not have PF *Mycobacterium* reads whereas some people without dTB have *Mycobacterium* reads, 6) CRP did not differ by TB status or HIV status, and α-diversity was similar by CRP status, however, β-diversity differed, with *Pseudomonas* enrichment in people with low (vs. high) CRP. This is the first characterisations of the site-of-disease microbiome in people with pericarditis, specifically in a high TB HIV-endemic setting.

dTBs were enriched in several taxa in addition to *Mycobacterium*. These include *Kocuria*, a skin commensal that causes an infection of the heart e.g., native valve endocarditis, albeit rarely^31^, and *Weeksella* which can also cause serious bacterial infections like bacteraemia, and spontaneous bacterial peritonitis^32^. The relevance of these taxa in TBP requires further investigation.

*Mycobacterium* was enriched in PLHIV compared to people without HIV. Furthermore, *Bifidobacterium* was associated with clinical features characteristic of severe disease (HIV-positivity and, in those with HIV, no ART and lower CD4 counts. *Bifidobacterium*, while commonly considered a beneficial, immunomodulatory gut commensal^33^, has been described as either depleted^9^ ^34^ or enriched^35^ ^36^ in people with pulmonary TB compared to symptomatic controls. However, our findings are consistent with the latter. *Bifidobacterium* can act opportunistically, causing bacteraemia in immunocompromised people and those with a damaged intestinal barrier^37^.

Similar to previous work involving people with different forms of TB (i.e., pulmonary TB and TB lymphadenitis^38^), people with TBP were enriched in SCFA-producing bacteria (e.g., *Lacticigenium* produces acetic and lactic acid) and pathways (*L-lysine fermentation to acetate and butanoate*, amino acid and carbohydrate metabolism butyrate precursor pathways)^39^ ^40^. dTBs also had elevated PF concentrations of LDH, a marker of tissue damage, however we acknowledge that it could include microbially-derived LDH. Lactate metabolism is also important for *Mtb* growth^41^

Furthermore, HIV-positive dTBs not on ART are, in addition to *Mycobacterium*, enriched in *Bifidobacterium*, a known producer of lactate which itself is also an intermediate metabolite of the butyrate biosynthetic pathway^42^ ^43^. Butyrate suppresses protective host responses (IFN-γ and IL-17A) to *Mtb.* Future studies should evaluate whether enhanced site-of-disease SCFA production correlates with both local and peripheral inflammation.

Pericarditis is often associated with pulmonary parenchymal disease. Notably, the PF microbiome was associated with a greater probability of chest imaging characteristics where individuals with PI showed distinct PF β-diversity compared to those without, characterized by increased *Mycobacterium* (likely resulting from more direct contact with the route of infection and site of pulmonary disease) and reduced *Streptococcus*. Similarly, individuals with pleural effusion exhibited different PF β-diversity compared to those without, with higher levels of *Streptococcus* and lower levels of *Brachymonas*. Pulmonary infiltrates are commonly observed in radiological imaging of individuals with pulmonary TB^44^, while pleural effusions are predominantly caused by TB in developing countries^45^. Consequently, obtaining sputum cultures and Xpert tests is crucial in suspected cases of TBP, even when parenchymal involvement is not evident. Furthermore, the co-existence of these features may reflect the TB route from the lungs to the heart, and what taxa might accompany that spread.

People within each TB group had *Mtb* DNA detected. The reasons for this are likely multifaceted: primarily due to technical methodological limitations (metagenomic sequencing would have more discriminatory power for species within *Mycobacteria*) but could also be due to TB exposure, past infection, TB disease at another anatomical site, or cross-contamination. It is important to note that 66% of dTBs did not show *Mycobacterium* readings, likely due to the limited sensitivity of 16S rRNA sequencing for detecting this genus, whose cells often lyse and release DNA inefficiently and may be biased against by conventional sequencing approaches (a “mycobacteriome” approach could be used to study mycobacterial diversity further)^46^.

CRP is a commonly studied biomarker in TB, and during active TB infection, CRP levels tend to be elevated due to the body’s immune response to the mycobacterial infection^47^. Similarly, CRP levels can be elevated in individuals with HIV because HIV infection is associated with chronic inflammation and immune activation, particularly in those with advanced disease or opportunistic infections^48-50^. However, in our study, unlike previously reported studies^51-53^, the level of CRP, appears to not be influenced by the presence of active TB in TBP. Despite this, the median CRP value in our TBP cohort 125.4 (70.83 – 176.70) ng/mL exceeded the diagnostic threshold ^54^ for TB diagnosis. This observation can be attributed to the fact our study only included people that were suspected to have TPB, leading to the presence of specific clinical symptoms and some form of the disease, regardless of whether it was TB or another condition. As a result, this could have contributed to the elevated CRP levels to some extent. Notably, we observed a depletion of *Pseudomonas* in presumptive TBP people with high CRP compared to those with low CRP, similar to a report of an adverse relationship between sputum genus richness and CRP in bronchiectasis^55^ ^56^.

The study has strengths and limitations. DNA-based sequencing was employed to analyze microbial communities, and alternative methods like culturomics, metagenomics, or metatranscriptomics may yield different results. Furthermore, as our study was nested within TB care at our hospital, our study data were influenced by limitations in routinely available data (e.g., unknown ART duration); however, this allowed us to recruit a high number of people. We also did background sampling and sequencing to detect potential cross-contamination of our low biomass fluid (PF), which allowed us to confirm that none of the discriminatory taxa identified in our analyses likely resulted from contamination. The study used a relaxed FDR-adjusted p-value threshold of 0.2 to identify various taxa, prioritizing hypothesis generation. This threshold was chosen as lower thresholds did not yield significant taxa, underscoring the need for broader exploration. We do not know which microbes are driving the high CRP in TB, however, this study was not designed to investigate this, and further studies would be required. We acknowledge the non-specificity of CRP as a general marker of inflammation. Finally, this study is a cross-sectional study, so it’s unable to establish temporal relationships.

In summary, the microbiomes of people with TBP (including those co-infected with HIV) are different to people without TBP and characterised by enriched *Mycobacterium* and other potentially immunomodulatory bacteria (*Bifidobacterium*, *Lacticigenium).* Further research is needed to define these mechanistic relationships (e.g., through murine models), including how the microbiota is altered by treatment and its association, if any, with clinical outcomes. This will provide insights into the development of effective host-directed therapies for TBP.

## Acknowledgments

The authors thank all study participants and CLIME staff, especially Sr Ruth Wilson. Computations were performed using facilities provided by the University of Cape Town’s ICTS High Performance Computing team: *<u>hpc.uct.ac.za</u>*.

## Data availability

Data and code will be made available upon publication. Other supporting data are available from the corresponding author (GT) on reasonable request.

