## Supplementary material for "Bad company? The pericardium microbiome in people investigated for tuberculosis pericarditis in an HIV-prevalent setting"

### **Methods**

#### *Aspiration procedure and pericardial fluid collection*

Samples were collected in the operating room under rigorous sterile conditions during echocardiogram-guided pericardiocentesis as follows:

1. The patient’s skin was cleaned with 0.5% chlorhexidine in 80% methylated spirit solution over the planned puncture site and allowed to dry.
2. Their skin was swabbed with an alcohol pad which was stored in a container with ~5ml sterile saline (**skin control**).
3. The patient was draped with sterile green surgical towels, leaving the puncture site covered with an adhesive sterile plastic Opsite dressing. The main purpose of this dressing was to fix and hold the sterile green towels in place as it adheres to both the skin and the edges of the green towels that have been positioned to expose the puncture site.
4. Two percent lignocaine was infiltrated into the skin and subcutaneous tissue at the intended site of puncture with a sterile syringe and needle.
5. The pericardium was punctured again with a separate sterile puncture needle attached to a sterile syringe filled with approximately 3ml of sterile saline.
6. The syringe was removed, and a 0.35-inch sterile J-tipped guidewire was advanced into the pericardial space.
7. The needle was removed, and an incision made into the skin with a sterile blade.
8. A 6-French sterile vascular sheath was flushed with sterile saline outside of the patient, and the washing was stored (**sheath control**).
9. The sheath was then advanced into the pericardial space over the guidewire.
10. The guidewire and dilator (within the sheath) were removed, leaving the sheath *in situ* in the pericardial space.
11. Pericardial fluid (PF) was collected with another sterile syringe directly from the sheath.

Control samples and PF were stored in the same specimen collection container types.

#### *Pericardial fluid tests*

Pericardial (fluid and/or tissue biopsy), non-pericardial (e.g., sputum showing the presence of proved tuberculosis elsewhere in the body), and blood specimens were sent to the National Health Laboratory Services (NHLS) for programmatic testing looking for microbiological evidence of TB other biomarkers in people with an inflammatory pericardial exudate. This included microbiology (*Mycobacterium tuberculosis* polymerase chain reaction testing, smear stain and Mycobacteria Growth Indicator Tube 960 (MGIT960) culture), PF and blood biochemistry (total protein, albumin, lactate dehydrogenase [LDH], adenosine deaminase [ADA]), cytology, haematology, histopathology, cytochemistry. Further, serological testing for C-reactive protein (CRP) and gamma interferon (IFN-γ) were performed.

#### *Definitions*

People were categorised based on extrapulmonary (PF and/or tissue biopsy, or non-pericardial (e.g., pleural fluid), or pulmonary (e.g., sputum) specimen mycobacteriological evidence, cytological evidence, and/or the clinical decision to start TB treatment by the attending clinician **(****Table S1)**. Definite tuberculous pericarditis (dTB) had at least one *Mycobacterium tuberculosis* complex (MTBC) positive (pericardial or non-pericardial) specimen by Mycobacteria Growth Indicator Tube (MGIT) 960 liquid culture (culture; BD Diagnostics), Xpert MTB/RIF (Xpert; Cepheid) and/or Xpert MTB/RIF Ultra (Ultra), or acid‐fast bacilli (AFB) staining microscopy*.* Probable tuberculous pericarditis (pTB) did not meet dTB criteria but commenced treatment empirically based on clinical suspicion, and no alternative diagnosis known. Non-tuberculous pericarditis (nTB) people had no microbiological evidence of MTBC, had or did not have an alternative diagnosis, and were not placed on treatment.

*Microbiota analysis*

##### Environmental and background controls**:**

Given the concern for potential carry-over of skin commensals and DNA present on medical apparatus, PF microbiome readouts were analysed in parallel with those from skin (puncture site swab) and the saline flush of the sterile vascular sheath (background control, BKG). These were analysed in a paired manner for 1 in 5 participants. Using the *decontam^1^* package, analysis was done to identify possible contaminating ASVs to ensure that the microbial patterns described were associated with the signals of those found in the pericardium space, and not include possible contaminating signatures. The prevalence-based contaminant identification method was used to assess the mean relative abundance of ASVs from PF and the two controls, which is preferred for samples of low biomass. ASVs identified as possible contaminants (**Table S2**) were not removed but when, observed in downstream *DESeq2* analyses, greyed out in volcano plots to reflect their status as possible contaminants.

##### 16S rRNA gene sequencing analysis**:**

The proportions test was done using STATA (v16; StataCorp, USA) to determine whether a specific variable was more frequent in different groups (e.g., people of different case definitions)^2^. The Mann-Whitney or Wilcoxon signed rank test was used for unpaired and paired comparisons between two groups respectively (e.g., α-diversity). The Kruskal-Wallis or Friedman test was used for unpaired and paired comparisons respectively, with Dunn’s test for multiple comparisons used for comparisons involving more than two groups. Spearman’s Rho rank correlation was used to measure the association between mycobacterial relative abundance and continuous variables. For analysis of continuous variables in different groups (e.g., PF size different TB status), the D’Agostino-Pearson omnibus normality test was done to evaluate normality, and the relevant parametric or nonparametric test was chosen based on the normality test. Analyses were done in GraphPad Prism (v7; GraphPad Software, USA) and R (v4.2; R Core Team). To assess whether factors other than active TB influenced PF microbial composition, we performed multi-factor PERMANOVA analysis.

##### Clustering**:**

Dirichlet multinomial mixture modelling (DMM) was performed using the R package *DirichletMultinomial* to establish clustering within on PF samples ^3^. Using genus tables, the number of clusters was determined by selecting the number of Dirichlet components that reduced the Laplace approximation of the model^3^ (i.e. lower values indicate better fits). Clustering profiles indicate unique groupings.

### **Results**

#### *Cohort characteristics*

dTBs were more likely to have HIV, cough, weigh less, be using other non-TB medications, have lower Albumin levels, and have higher ADA, LDH, and uIFN-γ fluid levels than nTBs. pTBs were more likely to have HIV, have had TB previously, and were already on TB treatment (less than 2 weeks), more self-reported symptoms, have lower Albumin levels, and have higher ADA and uIFN-γ fluid levels, than nTBs. dTBs were less likely to already be on TB treatment at recruitment (< 2 weeks) compared to pTBs (**Table 1**).

#### *Environmental and background controls*

Given the concern for potential carry-over of skin commensals and DNA present on medical apparatus, PF microbiome readouts were analysed in parallel with those from skin (puncture site swab) and the saline flush of the sterile vascular sheath (background control, BKG). These were analysed in a paired manner for 1 in 5 participants. Using the *decontam^1^* package, analysis was done to identify possible contaminating ASVs to ensure that the microbial patterns described were associated with the signals of those found in the pericardium space, and not include possible contaminating signatures. The prevalence-based contaminant identification method was used to assess the mean relative abundance of ASVs from PF and the two controls, which is preferred for samples of low biomass. ASVs identified as possible contaminants (**Table S2**) were not removed but when, observed in downstream *DESeq2* analyses, greyed out in volcano plots to reflect their status as possible contaminants.

#### *α- and β-diversities according to demographic, clinical, and microbiological characteristics (Table S2)*

Overall: Serous PF had a higher α-diversity than pyopericardium PF (*p*=0.031). β–diversity differed by TB status (*p*=0.025), ART status (*p*=0.001), weight loss (*p*=0.042), between people that had pulmonary infiltrates (*p*=0.026), and pleural effusion (*p*=0.030) compared to those that didn’t, between serous and serous-sanguineous (*p*=0.021), and pyopericardium PF (*p*=0.007). Similar trends in TB groups were observed when people with antibiotic use within the last year were excluded.

dTBs: β–diversity differed by ART status (*p*=0.010), between people that had pulmonary infiltrates compared to those that didn’t (*p*=0.006), and between serous and serous-sanguineous PF (*p*=0.037).

pTBs: α-diversity was less in people that had fever (*p*=0.049), and experienced night sweats (*p*=0.046). β–diversity differed by ART status (*p*=0.024).

nTBs: α-diversity was less in blood-stained PF compared to serous PF (*p*=0.042).

Some people had commenced TB treatment, but for less than 2 weeks only, however, there was no difference in the microbial diversity of these individuals when they were compared to people not on TB treatment (**Table S2**).

Some people included in this study were on other non-TB antibiotics either at the time of recruitment or recently prior (recorded people on non-TB antibiotics within 1 year of recruitment). Even though certain non-TB antibiotics have antimicrobial action, e.g., augmentin and levofloxacin, our analyses showed that this did not differ in frequency and was not associated with any microbial diversity of compositions, thus unlikely a confounder in our cohort.

Multivariate PERMANOVA analysis did not show additional associations with β-diversity (**Table S3**) other than TB.

#### *Correlation between 16S rRNA gene sequencing and TB diagnostic test results*

As expected, dTBs had a higher relative abundance of *Mycobacterium* (0.0001 [0.0001-0.0061] vs. 0.0001 [0.001-0.001], *p<0.017;* **Figure 2F**]. *Mycobacterium* reads were present in 34% (31/91) of dTBs, 8% (2/25) of pTBs and 17% (4/23) of nTBs (**Figure S6**). *Mycobacterium* relative abundance in dTBs showed a positive correlation with bacillary load (based on Xpert and Ultra cycle threshold values; r=-0.361, *p<0.0117*; **Figure S7A**)*,* and MGIT960 culture days-to-positivity; r=-0.484, *p=0.0001*; **Figure S7B**).

#### *HIV is not associated with microbiome differences in high vs. low CRP presumptive TBPs*

CRP levels were similar between HIV-positives [130.40 (IQR 97.40 – 183.8) ng/mL] and HIV-negatives [100.40 (IQR 49.45 – 176.70) ng/mL] in presumptive TBP people (*p*=0.172; **Figure S13A**).

HIV-positives: The relative abundance of *Mycobacterium* was similar between HIV-positives with high CRP [1.000e-005 (IQR 1.000e-005 – 0.005)] vs. low CRP [1.000e-005 (1.000e-005 – 0.0006), *p*=0.475; **Figure S13B**). α- and β-diversity was similar between HIV-positives with high vs. low CRP (**Figure S13C-D**), where high CRP individuals were *Caulobacter*-, *Leptothrix*- and *Sphingobium*-enriched, and *Rothia*- and *Staphylococcus*-depleted vs. low CRP individuals (**Figure S13E**). α- and β-diversities were similar within TB groups in HIV-positives (**Table 2**).

HIV-negatives: The relative abundance of *Mycobacterium* was similar between HIV-negatives with high [1.000e-005 (IQR 1.000e-005 – 1.000e-005)] vs. low CRP [1.000e-005 (1.000e-005 – 1.000e-005), *p*>0.999; **Figure S14A**). α- and β-diversity was similar between HIV-negatives with high vs. low CRP (**Figure S14B-C**), where high CRP individuals were *Thermicanus*-, *Pseudoxanthomonas*- and *Sphingobium*-enriched, and *Pseudomonas*-, *Lacticigenium*- and *Pelomonas*-depleted vs. low CRP individuals (**Figure S14E**). α- and β-diversities were similar within TB groups in HIV-negatives (**Table 2**).

#### *Dirichlet multinomial mixtures modelling*

We then evaluated for presence of distinct microbial communities in PF using DMM on all pericardial fluid samples. There were no clusters that were found (**Figure S15**) suggesting that the no distinct profiles are found in the PF microbiome. We previously identified three lymphotypes in TB lymphadenitis (another form of EPTB) of which the *Mycobacterium*-dominated cluster correlated with clinical markers of severe disease (i.e., HIV status, CD4 count, lymph node size)^4^.

Table S1: Reference standard definition used in the study.

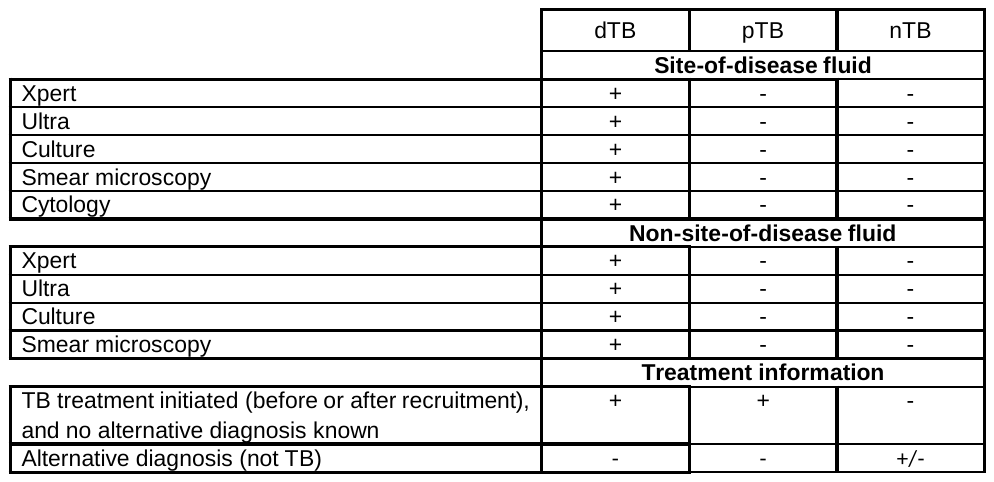
Abbreviations: dTB: definite tuberculous pericarditis; pTB: probable-tuberculous pericarditis; nTB: non-tuberculous pericarditis; Xpert: Xpert MTB/RIF; Ultra: Xpert MTB/RIF Ultra; MGIT960 Culture: Mycobacteria Growth Indicator Tube 960 liquid culture

Table S2: α- and β-diversities in people with presumptive TB pericarditis when people with different demographic and clinical characteristics were compared. Several characteristics, described in the Supplementary Results text, were associated with differing diversities.

| **Characteristics** | **Overall (n=139)** | | | **dTB (n=91)** | | | **pTB (n=30)** | | | **nTB (n=18)** | | |
| --- | --- | --- | --- | --- | --- | --- | --- | --- | --- | --- | --- | --- |
|  | α-diversity | β-diversity | | α-diversity | β-diversity | | α-diversity | β-diversity | | α-diversity | β-diversity | |
|  |  | *p-value* | *R*^2^ *value* |  | *p-value* | *R*^2^ *value* |  | *p-value* | *R*^2^ *value* |  | *p-value* | *R*^2^ *value* |
| TB | 0.874 | **0.025** | 0.022 | - | - | - | - | - | - | - | - | - |
| Sex | 0.733 | 0.162 | 0.009 | 0.529 | 0.599 | 0.010 | 0.657 | 0.632 | 0.036 | 0.242 | 0.309 | 0.049 |
| Current tobacco smoker | 0.628 | 0.127 | 0.009 | 0.769 | 0.289 | 0.012 | 0.484 | 0.709 | 0.034 | 0.142 | 0.315 | 0.049 |
| Previous tobacco smoker | 0.396 | 0.877 | 0.007 | 0.443 | 0.885 | 0.010 | 0.933 | 0.731 | 0.046 | 0.313 | 0.054 | 0.097 |
| Clinical characteristics: | | | | | | | | | | | | |
| HIV | 0.594 | 0.222 | 0.009 | 0.454 | 0.153 | 0.014 | 0.634 | 0.702 | 0.034 | - | - | - |
| On ART | 0.093 | **0.001** | 0.041 | 0.293 | **0.010** | 0.041 | 0.135 | **0.024** | 0.106 | - | - | - |
| Previous TB | 0.344 | 0.188 | 0.009 | 0.706 | 0.574 | 0.010 | 0.213 | 0.348 | 0.043 | 0.651 | 0.646 | 0.038 |
| Self-reported symptoms: | | | | | | | | | | | | |
| Cough | 0.930 | 0.368 | 0.008 | 0.576 | 0.151 | 0.015 | 0.391 | 0.612 | 0.038 | 0.949 | 0.273 | 0.051 |
| Fever | 0.083 | 0.219 | 0.009 | 0.422 | 0.519 | 0.011 | **0.049** | 0.167 | 0.056 | 0.766 | 0.492 | 0.043 |
| Night sweats | 0.361 | 0.135 | 0.010 | 0.785 | 0.417 | 0.012 | **0.046** | 0.333 | 0.045 | 0.606 | 0.120 | 0.059 |
| Weight loss | 0.493 | **0.042** | 0.012 | 0.521 | 0.298 | 0.013 | 0.975 | 0.976 | 0.027 | 0.606 | 0.138 | 0.059 |
| Chest X-ray results: | | | | | | | | | | | | |
| Normal | 0.171 | 0.730 | 0.006 | 0.234 | 0.766 | 0.009 | - | - | - | 0.585 | 0.178 | 0.059 |
| Cardiomegaly | 0.201 | 0.056 | 0.011 | 0.151 | 0.455 | 0.011 | 0.641 | 0.051 | 0.065 | 0.699 | 0.121 | 0.059 |
| Pulmonary infiltrates | 0.336 | **0.026** | 0.012 | 0.590 | **0.006** | 0.024 | 0.159 | 0.214 | 0.051 | 0.627 | 0.999 | 0.023 |
| Hilar lymphadenopathy | 0.815 | 0.670 | 0.006 | 0.582 | 0.720 | 0.009 | 0.133 | 0.323 | 0.049 | 0.663 | 0.406 | 0.046 |
| Miliary pattern | 0.610 | 0.153 | 0.009 | 0.598 | 0.253 | 0.013 | >0.999 | **0.038** | 0.052 | - | - | - |
| Pleural effusion | 0.606 | **0.030** | 0.012 | 0.920 | 0.133 | 0.015 | 0.821 | 0.220 | 0.048 | 0.176 | 0.450 | 0.044 |
| Pericardial tamponade | 0.421 | 0.387 | 0.008 | 0.636 | 0.200 | 0.015 | 0.257 | 0.762 | 0.036 | 0.900 | 0.157 | 0.057 |
| Treatment history: | | | | | | | | | | | | |
| Current TB treatment (<2 weeks) | 0.553 | 0.205 | 0.009 | 0.778 | 0.307 | 0.012 | 0.365 | 0.693 | 0.034 | - | - | - |
| Other medications | 0.498 | 0.439 | 0.008 | 0.696 | 0.896 | 0.008 | 0.208 | 0.282 | 0.050 | 0.593 | 0.743 | 0.037 |
| Antibiotic use within 1 year | 0.890 | 0.314 | 0.009 | 0.484 | 0.207 | 0.015 | 0.634 | 0.636 | 0.037 | 0.231 | 0.577 | 0.045 |
| Current antibiotic use | 0.179 | 0.270 | 0.056 | 0.390 | 0.227 | 0.098 | 0.245 | 0.195 | 0.221 | - | - | - |
| Fluid characteristic (*vs. serous)*: | | | | | | | | | | | | |
| Blood-stained | 0.912 | 0.062 | 0.014 | 0.372 | 0.102 | 0.021 | 0.914 | 0.149 | 0.080 | **0.025** | 0.818 | 0.041 |
| Serous-sanguineous | 0.489 | **0.021** | 0.027 | 0.413 | **0.037** | 0.040 | 0.200 | 0.311 | 0.101 | 0.221 | 0.834 | 0.082 |
| Pyopericardium | **0.031** | **0.007** | 0.051 | 0.700 | 0.236 | 0.048 | 0.121 | 0.105 | 0.215 | - | - | - |
| Chylous | 0.859 | 0.058 | 0.038 | 0.782 | **0.038** | 0.065 | - | - | - | - | - | - |
| Patient demised | 0.501 | 0.085 | 0.011 | 0.418 | 0.079 | 0.017 | 0.439 | 0.663 | 0.036 | 0.417 | 0.659 | 0.041 |

**R*^2^ provides the proportion of variation explained (e.g., a factor that has a *R*^2^ = 0.037, explains 3.7% of the variation in community composition) by β-diversity. Abbreviations: ART: antiretroviral therapy; dTB: definite tuberculous pericarditis; nTB: non-tuberculous pericarditis; pTB: probable tuberculous pericarditis; TB: tuberculosis; TBP: tuberculous pericarditis.

Table S3: PERMANOVA of the Bray-Curtis dissimilarities for bacterial community structure used for detection of possible confounding variables associated with PTB. TB remained associated with differences in PF microbial composition in presumptive TBP people when other factors were included.

| **Characteristics** | ***R*^2^ *value*** | ***p*-value** |
| --- | --- | --- |
| TB | 0.02259 | **0.011** |
| Age, years | 0.35789 | 0.069 |
| Sex (male vs. Female) | 0.0072 | 0.236 |
| Current tobacco smoker | 0.0147 | 0.2 |
| Previous tobacco smoker | 0.00483 | 0.775 |
| HIV | 0.00882 | 0.079 |
| On ART | 0.00709 | 0.302 |
| CD4 count, cells/µl | 0.40563 | 0.099 |
| Previous TB | 0.01605 | 0.102 |
| Chest X-ray results: |  |  |
| Cardiomegaly | 0.00733 | 0.233 |
| Pulmonary infiltrates | 0.00525 | 0.667 |
| Hilar lymphadenopathy | 0.00707 | 0.239 |
| Miliary pattern | 0.0065 | 0.368 |
| Pleural effusion | 0.00537 | 0.645 |
| Antibiotic use within 1 year | 0.01635 | 0.097 |
| Fluid characteristics | 0.01364 | 0.28 |

**R*^2^ provides the proportion of variation explained (e.g., a factor that has a *R*^2^ = 0.037, explains 3.7% of the variation in community composition) by β-diversity. Abbreviations: ART: antiretroviral therapy; PF: pericardial fluid.

Figure S1: Environmental cross contamination is highly unlikely. (A) α-diversity analyses show PF has lower diversity than skin and BKG. (B) β-diversity of PF differs to BKG and skin. *DESeq2* volcano plots depicting differentially abundant taxa show that PF was enriched in (C) *Mycobacterium* vs. BKG and enriched in (D) *Mycobacterium* and *Streptococcus* vs. skin (red dotted line represents adjusted *p*-value=0.2; circle size represents relative abundance; taxa identified as possible contaminants are shown in grey). PF: pericardial fluid.

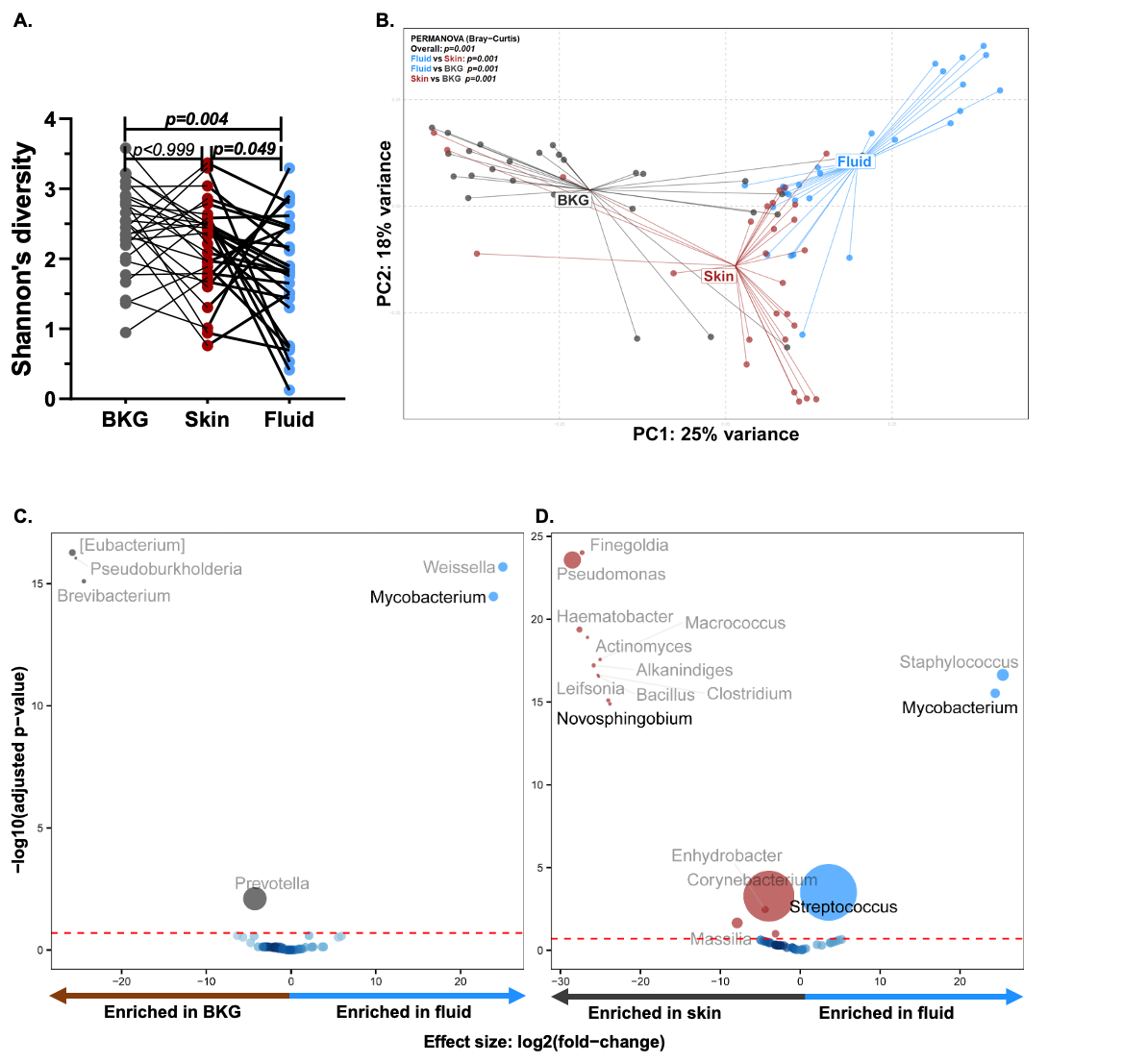

Figure S2: Relative abundances of the top 100 potential contaminating (pink) taxa in pericardial fluid vs. BKG based on the decontam prevalence method. Mean relative abundances of taxa in (A) BKG, (B) pericardial fluid, and (C) skin, are ranked by BKG in decreasing order.

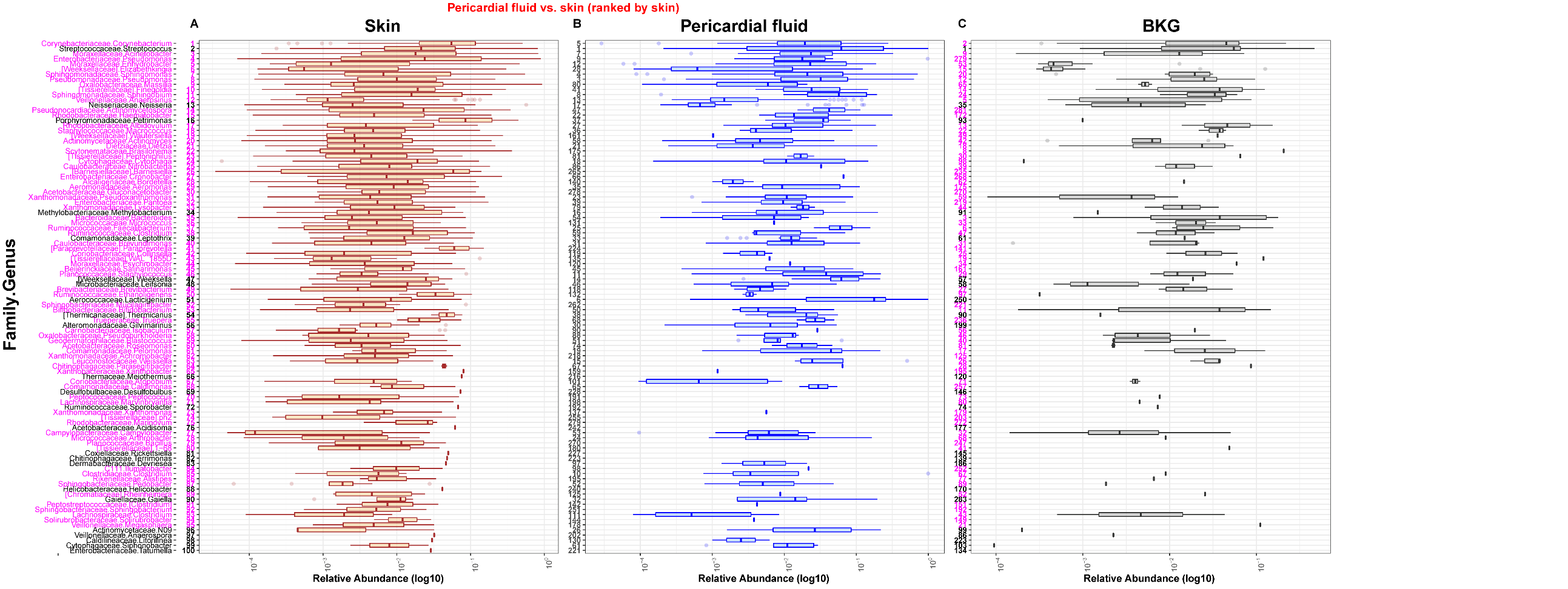
Figure S3: Relative abundances of the top 100 potential contaminating (pink) taxa in pericardial fluid vs. skin based on the decontam prevalence method. Mean relative abundances of taxa in (A) skin, (B) pericardial fluid, and (C) BKG, are ranked by skin in decreasing order.

Figure S4: PF microbiota of dTBs is more compositionally similar to each other than to pTBs and nTBs. Within-subject Bray dissimilarity distances were highest in pTBs, followed by nTBs, and lastly dTBs. dTB: definite tuberculous pericarditis; nTB: non-tuberculous pericarditis; PF: pericardial fluid; pTB: probable tuberculous pericarditis; TB: tuberculosis; TBP: tuberculous pericarditis.

Figure S5: Grouping pTBs with dTBs or nTBs resulted in no diversity differences. No α- or β-diversity differences were observed when pTBs were grouped with either (A-B) dTBs or (C-D) nTBs. (E-F) TBs were enriched with *Mycobacterium* and *Lacticigenium* versus non-TBs regardless of whom the pTBs were grouped with (red dotted line represents adjusted *p*-value=0.2; circle size represents relative abundance; taxa identified as possible contaminants are shown in grey). dTB: definite tuberculous pericarditis; nTB: non-tuberculous pericarditis; pTB: probable tuberculous pericarditis; TB: tuberculosis; TBP: tuberculous pericarditis.

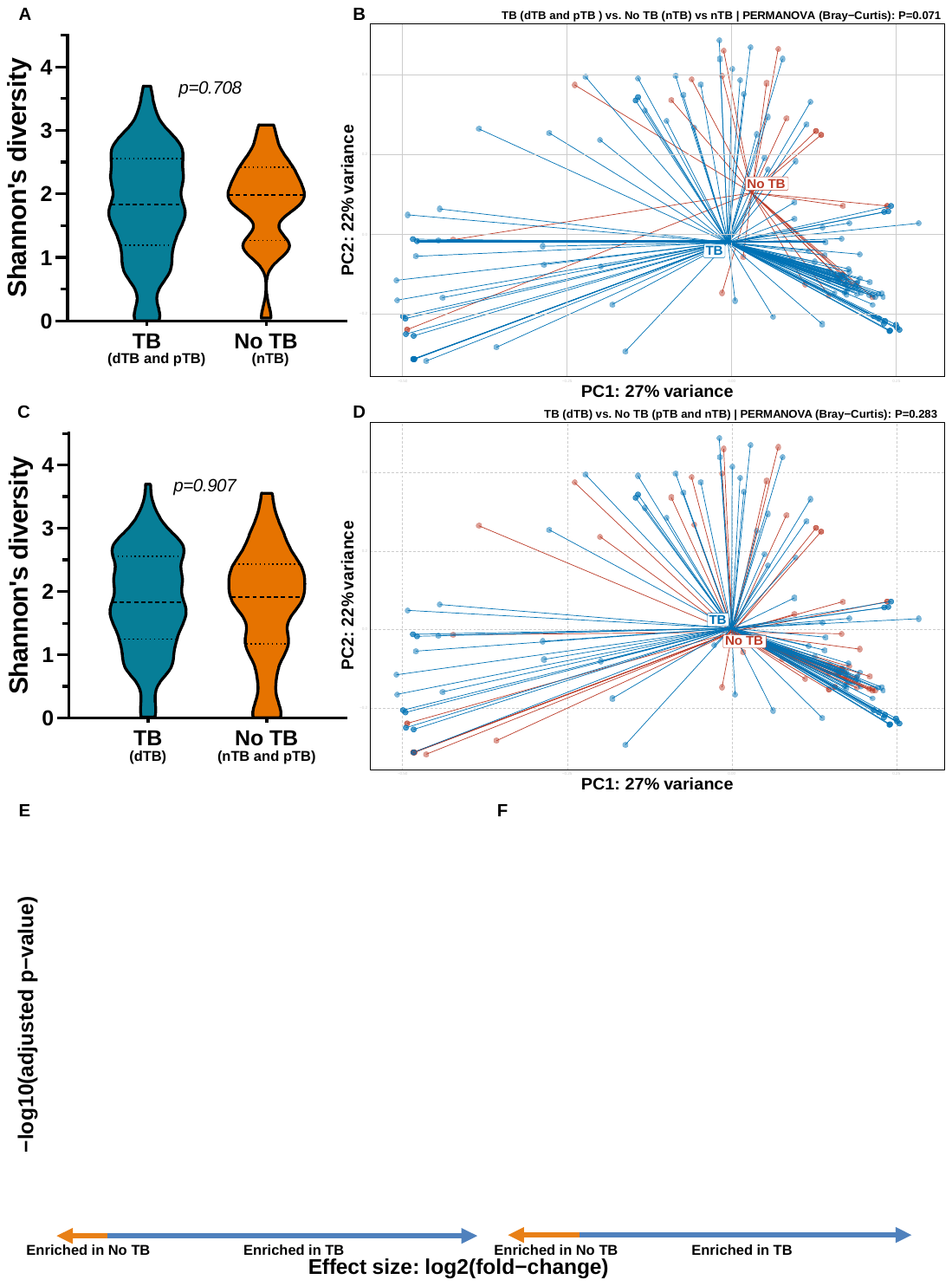

Figure S6: *Mycobacterium* reads per participant stratified by TB status shows *Mycobacterium* in some pTBs and nTBs. Furthermore, not all dTBs had detected *Mycobacterium* reads. dTB: definite tuberculous pericarditis; nTB: non-tuberculous pericarditis; pTB: probable tuberculous pericarditis; TB: tuberculosis; TBP: tuberculous pericarditis.

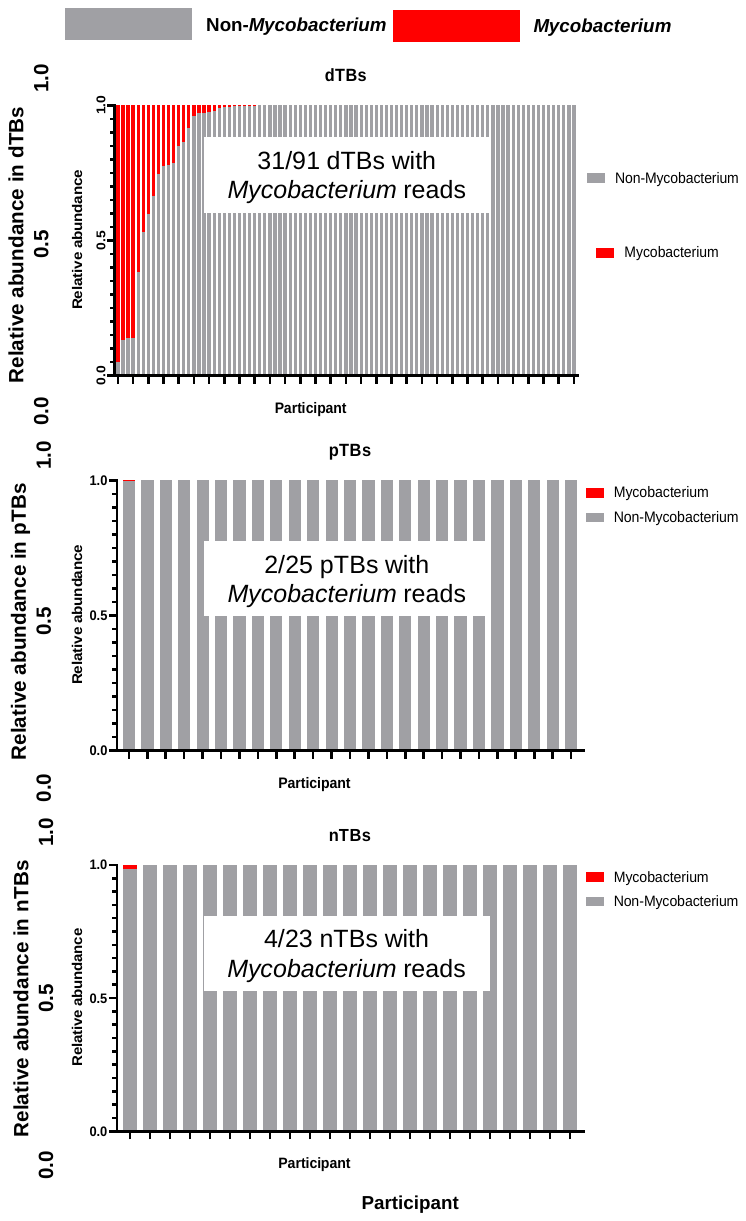

Figure S7: Pericardial fluid 16S rRNA gene sequencing is positively correlated with mycobacterial load from test results. In dTBs, relative abundances of mycobacterial reads present in the PF negatively correlated with (A) culture days-to-positivity, and (B) Xpert and Ultra mycobacterial load. Lower values indicate more mycobacteria. dTB: definite tuberculous pericarditis; PF: pericardial fluid; Xpert: Xpert MTB/RIF; Ultra: Xpert MTB/RIF Ultra; r_s_: Spearman correlation coefficient.

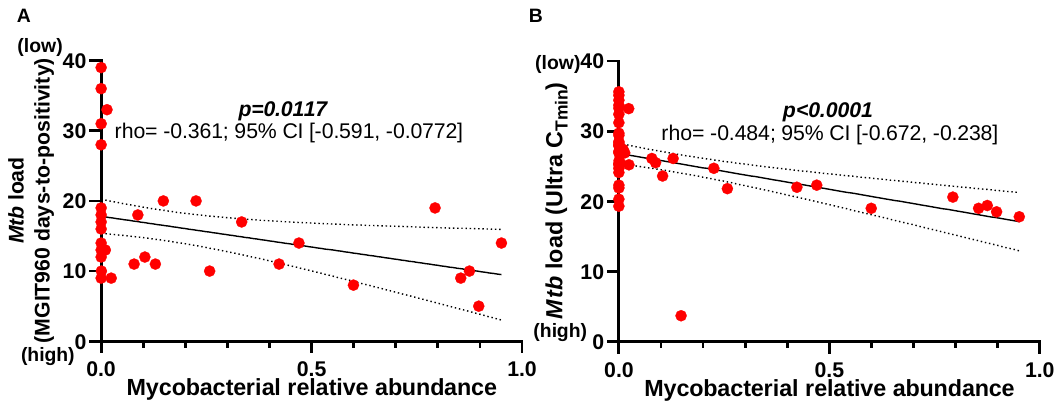

Figure S8: HIV is associated with the PF microbiota only in people co-infected with TB. In all people, (A) α- and (B) β-diversities did not differ by HIV status (solid circles: dTB; empty squares: pTB; empty circles: nTB). However, (C) *Mycobacterium* was enriched in HIV-positive compared to HIV-negative people. In dTBs, (D) *Mycobacterium*, *Bifidobacterium*, and *Leptothrix* were enriched in HIV-positive compared to HIV-negative people overall (red dotted line represents adjusted *p*-value=0.2; circle size represents relative abundance; taxa identified as possible contaminants are shown in grey). dTB: definite tuberculous pericarditis; HIV+: HIV-positive; HIV-: HIV-negative; nTB: non-tuberculous pericarditis; PF: pericardial fluid; pTB: probable tuberculous pericarditis; TB: tuberculosis; TBP: tuberculous pericarditis.

Figure S9: Microbiome composition variations in PLHIV and dTB cases are influenced by CD4 count and ART. (A) *Mycobacterium* is depleted in people with a high CD4 count in all PLHIV, and (B) dTBs. (C) PLHIV on ART were enriched in *Streptococcus* and depleted in *Mycobacterium* and *Bifidobacterium*. (D) dTBs on ART were depleted in *Mycobacterium* and *Bifidobacterium* whereas (red dotted line represents adjusted *p*-value=0.2; circle size represents relative abundance; taxa identified as possible contaminants are shown in grey). ART: antiretroviral therapy; dTB: definite tuberculous pericarditis; PLHIV: people living with HIV; pTB: probable tuberculous pericarditis; TB: tuberculosis; TBP: tuberculous pericarditis.

Figure S10: HIV-positive dTBs are depleted in biosynthesis pathways. Multiple pathways were depleted in HIV-positive dTBs compared to HIV-positive dTBs, all which are related to microbial biosynthesis. Red dotted line represents adjusted *p*-value=0.05; circle size represents relative abundance; pathways of interest are bolded. dTB: definite tuberculous pericarditis; HIV+: HIV-positive; HIV-: HIV-negative; TB: tuberculosis; TBP: tuberculous pericarditis.

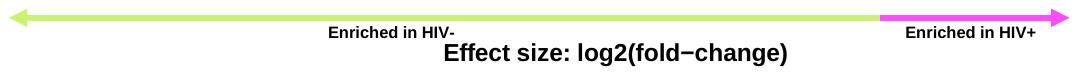

Figure S11: Intermediate SCFA metabolic pathways are enriched in dTBs. Among the pathways enriched in dTBs compared to pTBs were butyrate precursor pathways (amino acid and carbohydrate metabolism pathways). Red dotted line represents adjusted *p*-value=0.05; circle size represents relative abundance; pathways of interest are bolded. dTB: definite tuberculous pericarditis; pTB: probable tuberculous pericarditis; TB: tuberculosis; TBP: tuberculous pericarditis.

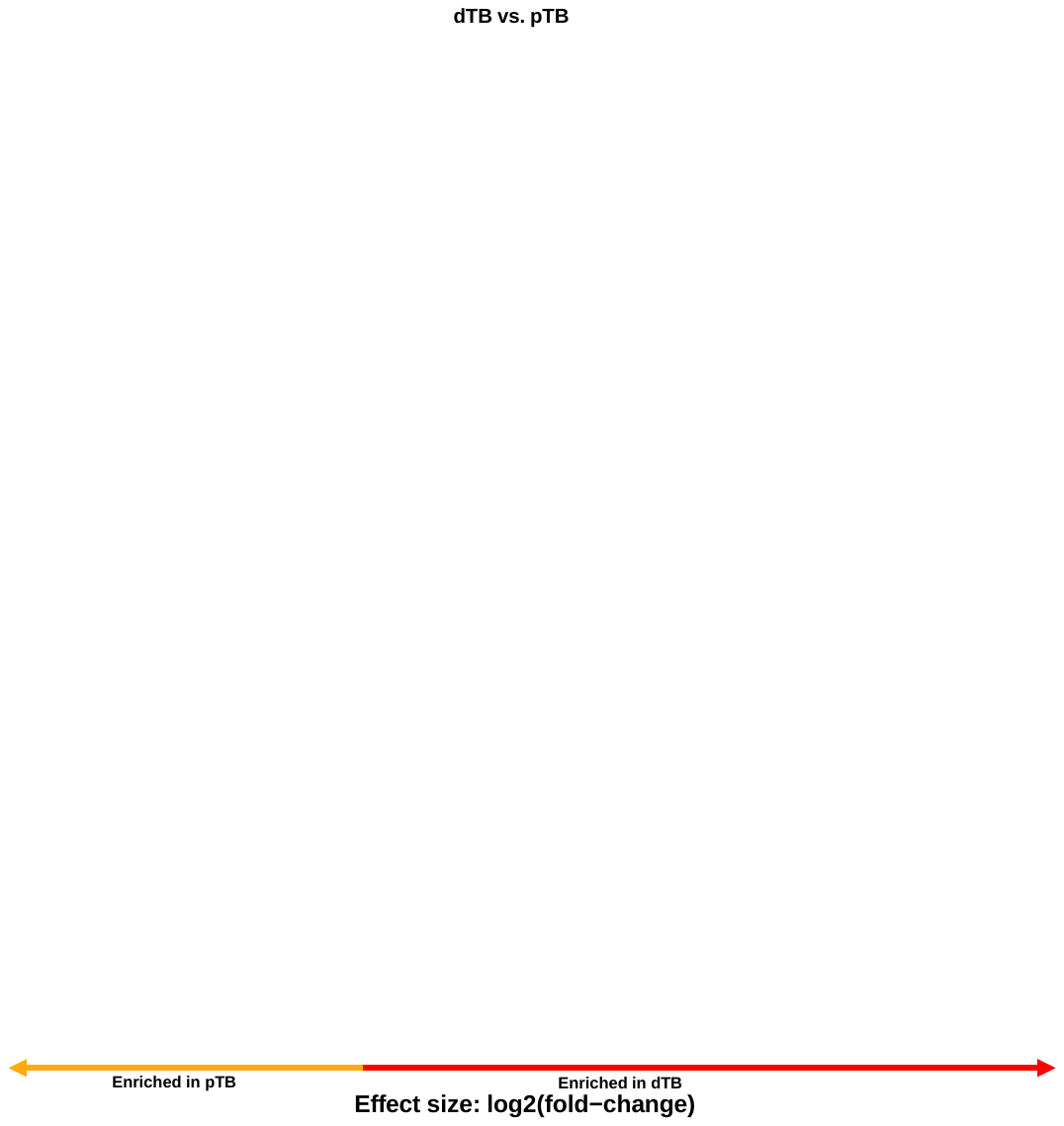

Figure S12: Enrichment of amino acid biosynthesis pathway in nTBs versus pTBs. nTBs had more amino acid biosynthesis pathways than pTBs. Red dotted line represents adjusted *p*-value=0.05; circle size represents relative abundance; pathways of interest are bolded. dTB: definite tuberculous pericarditis; nTB: non-tuberculous pericarditis; pTB: probable tuberculous pericarditis; TB: tuberculosis; TBP: tuberculous pericarditis.

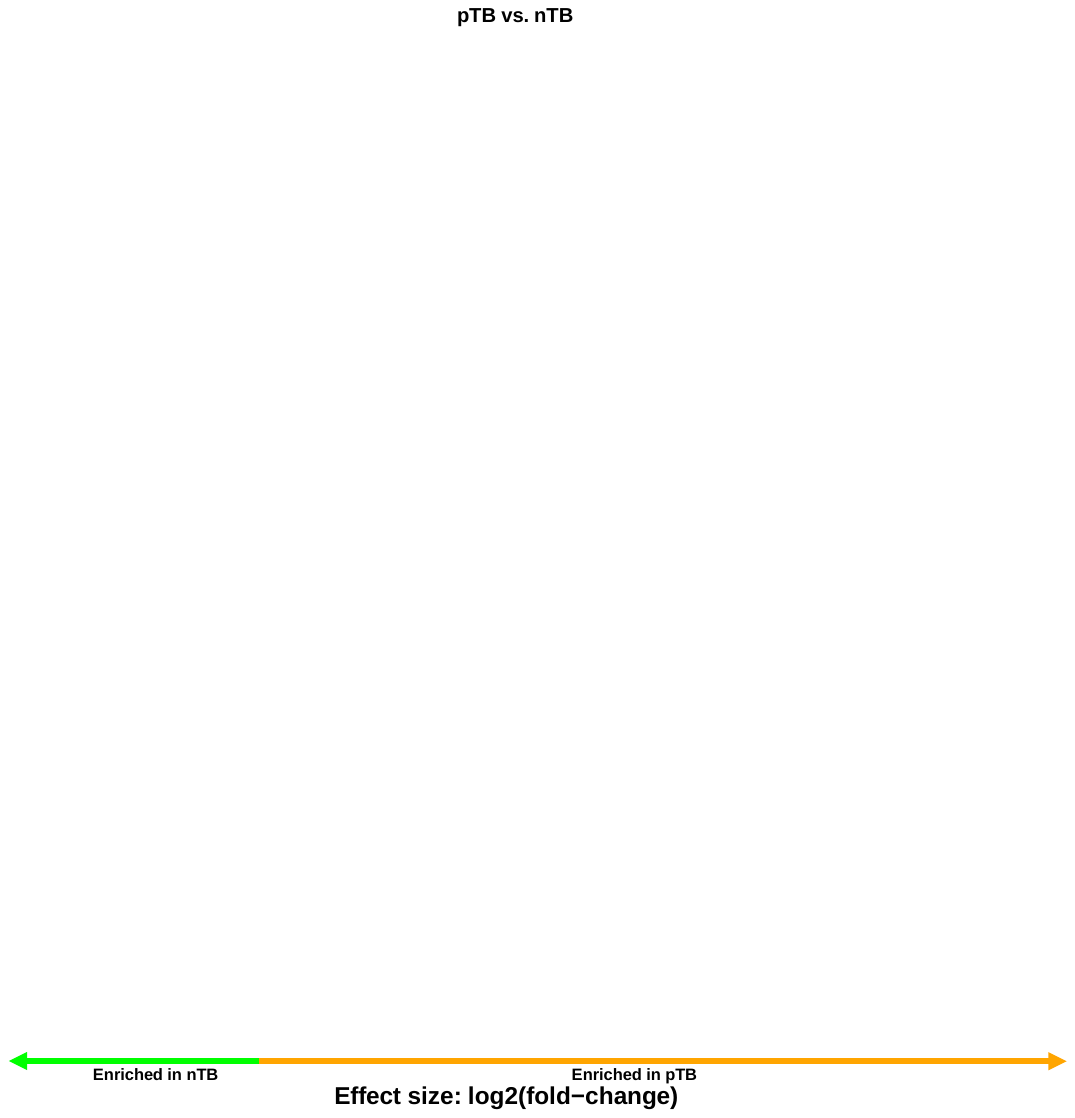

Figure S13: Microbime in HIV-positive TBP individuals with high CRP levels is similar compared to low CRP individuals. A) CRP levels showed no significant difference between HIV-positive vs. negative individuals with presumptive TBP. B) The relative abundance of *Mycobacterium* was comparable between HIV-positive individuals with high CRP. C) α- and D) β-diversity remained similar between HIV-positive individuals with high and low CRP. E) Among those with high CRP, there was an enrichment of *Caulobacter*, *Leptothrix*, and *Sphingobium*, while *Rothia* and *Staphylococcus* were depleted compared to low CRP individuals. Discriminatory taxa appear above the threshold (red dotted line, FDR=0.2); circle size represents relative abundance. CRP, C-reactive protein; FDR, false discovery rate; PERMANOVA, permutational multivariate analysis of variance; TBP: tuberculous pericarditis.

Figure S14: Mycobacterium relative abundance and microbiome is similar in HIV-negative TBP individuals with high vs. low CRP levels. A) The relative abundance of *Mycobacterium* was comparable between HIV-negative individuals with high vs. low CRP. Furthermore, B) α- and C) β-diversity remained similar between HIV-negative individuals with high and low CRP. However, D) However, individuals with high CRP exhibited enrichment in *Thermicanus*, *Pseudoxanthomonas*, and *Sphingobium*, while *Pseudomonas*, *Lacticigenium*, and *Pelomonas* were depleted compared to low CRP individuals. Discriminatory taxa appear above the threshold (red dotted line, FDR=0.2); circle size represents relative abundance. CRP, C-reactive protein; FDR, false discovery rate; PERMANOVA, permutational multivariate analysis of variance; TBP: tuberculous pericarditis.

Figure S15: No clusters were found when DMM was applied to all people. Laplace approximation predicts no clustering in pericardial fluid. Lower values indicate better fit.
